# Changes COVID-19 Post-Quarantine Behaviors, Hygiene and Expectations in Colombia: Population Survey from 1st to 13th September, 2020

**DOI:** 10.1101/2020.11.26.20239442

**Authors:** Sandra Marcela Sánchez, Danny Rivera-Montero, Rocío Murad-Rivera, Mariana Calderón-Jaramillo, Daniela Roldán, Lina María Castaño, Juan Carlos Rivillas-García

**Author notes:** The survey to collect data regarding COVID19 response in Colombia was executed based on a previous survey carried out on April 8, 2020, which was translated into Spanish and adapted with the consent of the School of Public Health at Imperial College London. Their module on Changes and Expectations was translated into Spanish and adapted with the consent of the Policy Institute at King’s College London. We express our gratitude to Professors Helen Ward and Christina Atkinson (Imperial’s SPH) and Bobby Duffy George Murkin (KCL) leader researchers and for allowing us to use their collection instrument.

## Abstract

This study by Profamilia Association focuses on the social response to COVID19 by reporting and analyzing the answers to two surveys carried out between April 16 and 25, and throughout September 2020. The study aims to identify changes in behaviors and immediate expectations after the quarantine was lifted. In general, results show that people have adopted behavioral changes such as wearing face masks, avoiding people with symptoms, and reducing mobility. However, it also shows that people’s concerns have doubled for many reasons, ranging from mental health issues, neglected sexual and reproductive health needs, the burden of care for others, and working from home. Similarly, it was found that some people would accept significant long-term changes such as accepting most children continuing to be homeschooled or employees choosing whether to work or not, targeted quarantines in neighborhoods that show high number of cases; and making the use of face masks mandatory in case a vaccine or treatment for Covid-19 does not become available. This means that during the quarantine and compared to April, many people consider these options to be acceptable in the long term.

The survey was filled out online via SurveyMonkey by 1,735 people in Colombia between 1st and 11th September, 2020. Overall, 17% mentioned that they had participated in the previous survey, Estudio Solidaridad I early stage of quarantine (April 2020). The following is a summary of the main findings based on the comparison of the two surveys in hopes to show evidence for changes in behavior, hygiene, levels of measure compliance, unmet needs, and to show peopleś immediate expectations after six months of strict quarantine.

- The most common concerns among the findings were: a vaccine or treatment for COVID-19 not arriving Colombia soon enough (79%), a vaccine or treatment not being developed soon enough (79%) and also a concern that once the vaccine arrives in Colombia, it will not be accessible (74%). 50% of people think it is likely to get the COVID-19 vaccine once it becomes readily available.
- 62% get information about COVID-19 through social media, 55% through official websites, and 51% through television.
- 43% say that in their neighborhood, community, social group, or town, measures and campaigns have been carried out to prevent the spread of the COVID-19.
- 25% say they would like to support local communities respond to the outbreak.
- 82% agree with most children remaining home-schooled.
- 85% think parents should be able to choose whether or not to send their children to school.
- 95% agree that people should be forced to wear face masks outside the home.
- 90% agree with neighborhoods, districts, or municipalities which experience outbreaks adopting more restrictive measures compared to the country as a whole.
- 86% think employees should choose whether to work in their office or work from home.
- 44% think that people will be able to be vaccinated against COVID-19 in a year or a year and a half.
- 26% think life will return to “normal” in two years or more.

## Introduction

An online survey was conducted in Colombia between April 8 and 20, 2020, and it was filled out by 3549 adults. This survey analyzed individuals’ social response to the measures adopted by the government to control COVID-19 regarding three different areas. Behaviors of hygiene, physical distancing such as a reduction in mobility and the ability to engage in mandatory and voluntary quarantines, and respondent perceptions of risk and ability to protect oneself and others (1); Unmet sexual and reproductive health needs (2); and the impact on mental health, and respondents’ perception of the response by the government (3). This first assessment also identified that there are at least three groups of people in the country who respond differently to the pandemic: those who resist the situation (34%), those who suffer from it (26%), and those who accept it (40%) (4).

**Table 1.**
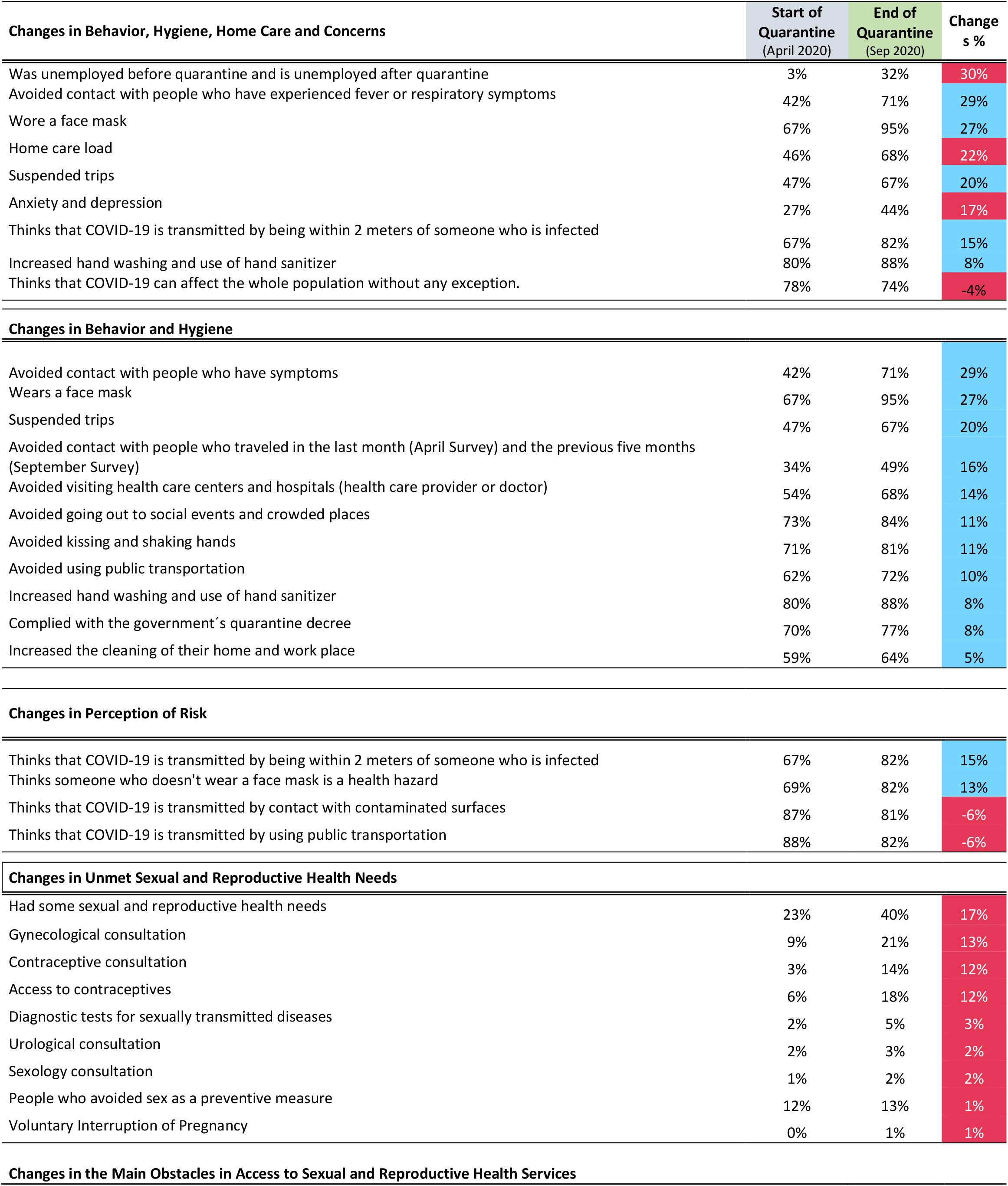
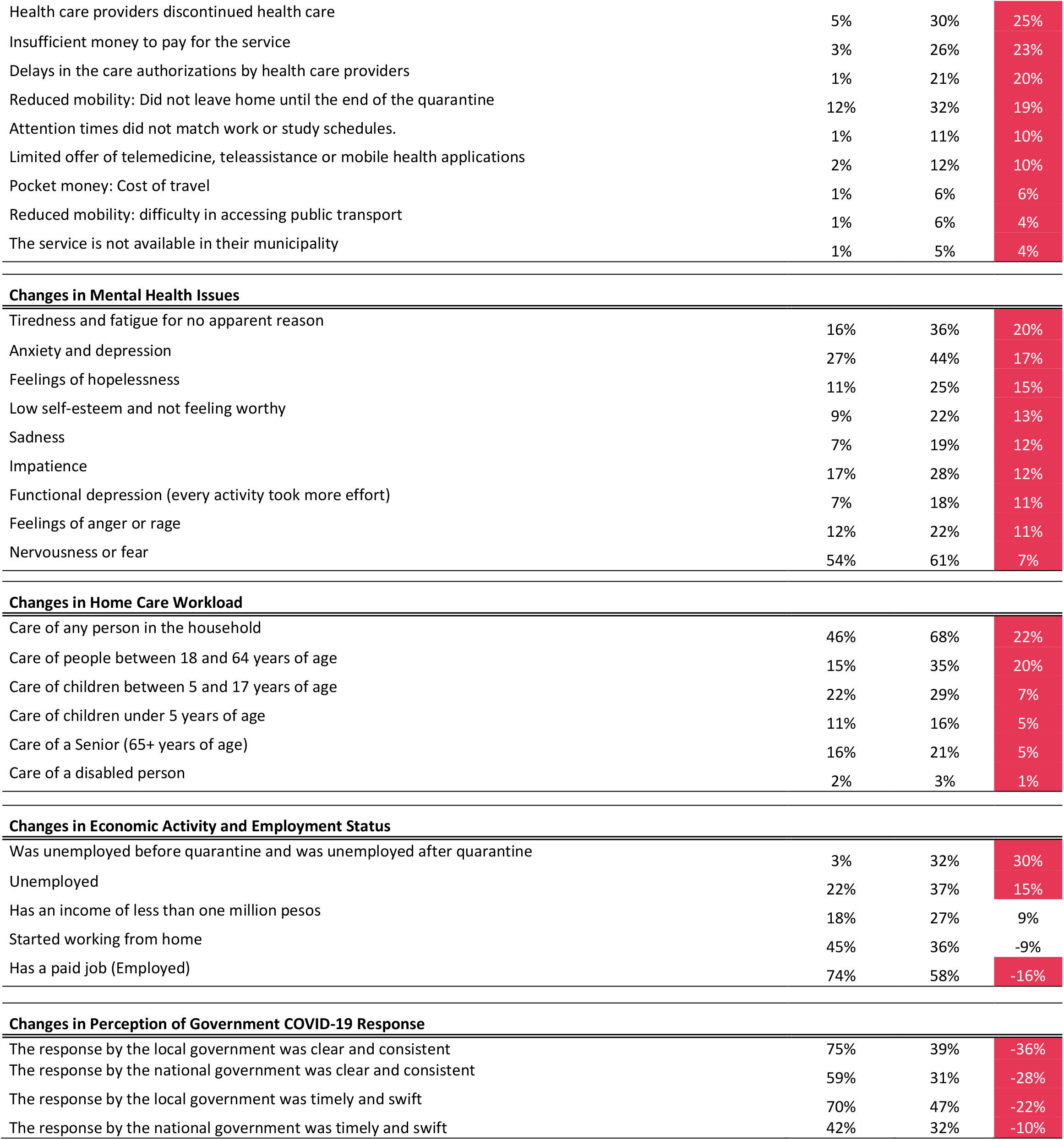
Changes in behavior, hygiene, home care and concerns, from the beginning of mandatory quarantine to the relaxation of measures.

| **Changes in Behavior, Hygiene, Home Care and Concerns** | **Start of Quarantine**<br>**(April 2020)** | **End of Quarantine**<br>**(Sep 2020)** | **Changes %** |
| --- | --- | --- | --- |
| Was unemployed before quarantine and is unemployed after quarantine | 3% | 32% | 30% |
| Avoided contact with people who have experienced fever or respiratory symptoms | 42% | 71% | 29% |
| Wore a face mask | 67% | 95% | 27% |
| Home care load | 46% | 68% | 22% |
| Suspended trips | 47% | 67% | 20% |
| Anxiety and depression | 27% | 44% | 17% |
| Thinks that COVID-19 is transmitted by being within 2 meters of someone who is infected | 67% | 82% | 15% |
| Increased hand washing and use of hand sanitizer | 80% | 88% | 8% |
| Thinks that COVID-19 can affect the whole population without any exception. | 78% | 74% | -4% |
| **Changes in Behavior and Hygiene** |  |  |  |
| Avoided contact with people who have symptoms | 42% | 71% | 29% |
| Wears a face mask | 67% | 95% | 27% |
| Suspended trips | 47% | 67% | 20% |
| Avoided contact with people who traveled in the last month (April Survey) and the previous five months (September Survey) | 34% | 49% | 16% |
| Avoided visiting health care centers and hospitals (health care provider or doctor) | 54% | 68% | 14% |
| Avoided going out to social events and crowded places | 73% | 84% | 11% |
| Avoided kissing and shaking hands | 71% | 81% | 11% |
| Avoided using public transportation | 62% | 72% | 10% |
| Increased hand washing and use of hand sanitizer | 80% | 88% | 8% |
| Complied with the government's quarantine decree | 70% | 77% | 8% |
| Increased the cleaning of their home and work place | 59% | 64% | 5% |
| **Changes in Perception of Risk** |  |  |  |
| Thinks that COVID-19 is transmitted by being within 2 meters of someone who is infected | 67% | 82% | 15% |
| Thinks someone who doesn't wear a face mask is a health hazard | 69% | 82% | 13% |
| Thinks that COVID-19 is transmitted by contact with contaminated surfaces | 87% | 81% | -6% |
| Thinks that COVID-19 is transmitted by using public transportation | 88% | 82% | -6% |
| **Changes in Unmet Sexual and Reproductive Health Needs** |  |  |  |
| Had some sexual and reproductive health needs | 23% | 40% | 17% |
| Gynecological consultation | 9% | 21% | 13% |
| Contraceptive consultation | 3% | 14% | 12% |
| Access to contraceptives | 6% | 18% | 12% |
| Diagnostic tests for sexually transmitted diseases | 2% | 5% | 3% |
| Urological consultation | 2% | 3% | 2% |
| Sexology consultation | 1% | 2% | 2% |
| People who avoided sex as a preventive measure | 12% | 13% | 1% |
| Voluntary Interruption of Pregnancy | 0% | 1% | 1% |

**Changes in the Main Obstacles in Access to Sexual and Reproductive Health Services**
| **Changes in Behavior, Hygiene, Home Care and Concerns** | **Start of Quarantine**<br>**(April 2020)** | **End of Quarantine**<br>**(Sep 2020)** | **Changes %** |
| --- | --- | --- | --- |
| Health care providers discontinued health care | 5% | 30% | 25% |
| Insufficient money to pay for the service | 3% | 26% | 23% |
| Delays in the care authorizations by health care providers | 1% | 21% | 20% |
| Reduced mobility: Did not leave home until the end of the quarantine | 12% | 32% | 19% |
| Attention times did not match work or study schedules. | 1% | 11% | 10% |
| Limited offer of telemedicine, teleassistance or mobile health applications | 2% | 12% | 10% |
| Pocket money: Cost of travel | 1% | 6% | 6% |
| Reduced mobility: difficulty in accessing public transport | 1% | 6% | 4% |
| The service is not available in their municipality | 1% | 5% | 4% |
| **Changes in Mental Health Issues** |  |  |  |
| Tiredness and fatigue for no apparent reason | 16% | 36% | 20% |
| Anxiety and depression | 27% | 44% | 17% |
| Feelings of hopelessness | 11% | 25% | 15% |
| Low self-esteem and not feeling worthy | 9% | 22% | 13% |
| Sadness | 7% | 19% | 12% |
| Impatience | 17% | 28% | 12% |
| Functional depression (every activity took more effort) | 7% | 18% | 11% |
| Feelings of anger or rage | 12% | 22% | 11% |
| Nervousness or fear | 54% | 61% | 7% |
| **Changes in Home Care Workload** |  |  |  |
| Care of any person in the household | 46% | 68% | 22% |
| Care of people between 18 and 64 years of age | 15% | 35% | 20% |
| Care of children between 5 and 17 years of age | 22% | 29% | 7% |
| Care of children under 5 years of age | 11% | 16% | 5% |
| Care of a Senior (65+ years of age) | 16% | 21% | 5% |
| Care of a disabled person | 2% | 3% | 1% |
| **Changes in Economic Activity and Employment Status** |  |  |  |
| Was unemployed before quarantine and was unemployed after quarantine | 3% | 32% | 30% |
| Unemployed | 22% | 37% | 15% |
| Has an income of less than one million pesos | 18% | 27% | 9% |
| Started working from home | 45% | 36% | -9% |
| Has a paid job (Employed) | 74% | 58% | -16% |
| **Changes in Perception of Government COVID-19 Response** |  |  |  |
| The response by the local government was clear and consistent | 75% | 39% | -36% |
| The response by the national government was clear and consistent | 59% | 31% | -28% |
| The response by the local government was timely and swift | 70% | 47% | -22% |
| The response by the national government was timely and swift | 42% | 32% | -10% |

Research in other countries has shown the importance of carrying out this type of study during gradual lifting or deconfinement periods, also known as the “relaxation” phase. The implications of lifting strict measures and the possibility for people to be released from quarantine require a series of physical distancing policies, alternative measures, testing, and efficient tracking of cases during, as well as considering the possibility of new outbreaks, even when COVID-19 is apparently under control (5-7).

Much of the physical distancing measures have started to be loosened in Colombia, and major cities are preparing for the so-called “new normal.” In the country, changes in behavior and immediate expectations during the national government’s relaxation phase are yet to be explored. Therefore, it is considered necessary and equally important to research changes in behavior, hygiene, compliance with physical distancing measures (mandatory and voluntary quarantine measures), and perceptions of risk and protection to care for oneself and others, unmet sexual and reproductive health needs, mental health effects, and perceptions of the government’s response to the pandemic. Again, this this new quarantine lifting phase offers a research opportunity to generate relevant and timely evidence to analyze behavioral changes to respond to COVID-19.

### General Objective

To analyze changes in behavior and immediate expectations during the relaxation and deconfinement phase decreed by the national government.

### Study Design and Survey Implementation

- Observational study based on a descriptive exploratory study.
- Total sample size was 1,735 adults (18 years and older) nationwide.
- The survey was conducted between September 1 and 11, 2020.
- The survey was conducted online using SurveyMonkey and was sent by email to contacts in Profamilia’s database and included partners and social networks (Twitter, Facebook, WhatsApp) and people who participated in the previous Solidaridad I survey.

## Methods

### Study and Sample Design

This research was a cross-sectional descriptive exploratory study. The online survey was developed on **SurveyMonkey®** and a non-probabilistic snowball sample was used to disseminate it because it allows the sample size to increase as the people initially selected to invite others to participate. The total number of completed surveys for analysis was 1,735. The survey was conducted in all departments of the country, except for Casanare, Vichada, Guainía, and Vaupés.

**Table 2.** Number of Surveys Conducted by Department.

| Department | Surveys conducted | Department | Surveys conducted |
| --- | --- | --- | --- |
| Antioquia | 341 | Meta | 10 |
| Atlántico | 75 | Nariño | 15 |
| Bogotá, D.C. | 593 | Norte de Santander | 37 |
| Bolívar | 30 | Quindío | 8 |
| Boyacá | 26 | Risaralda | 14 |
| Caldas | 17 | Santander | 56 |
| Caquetá | 3 | Sucre | 9 |
| Cauca | 21 | Tolima | 18 |
| Cesar | 3 | Valle del Cauca | 204 |
| Córdoba | 33 | Arauca | 5 |
| Cundinamarca | 113 | Putumayo | 1 |
| Chocó | 7 | San Andrés islands | 2 |
| Huila | 37 | Amazonas | 1 |
| La Guajira | 29 | Guaviare | 3 |
| Magdalena | 22 | NR | 2 |
|  |  | **Total** | **1.735** |

The survey in Colombia had six components: 1) socio-demographic characteristics; 2) care and employment burden of care; 3) perceptions of risk and protection; 4) behavior changes; 5) community mobilization and resilient practices; and 6) immediate changes and expectations.

Among the advantages of implementing an online survey through SurveyMonkey® is the speed and ease in which it is conducted as well as the opportunity it presented to take advantage of the quarantine, moment in which many households increased their connectivity and online presence. Among the limitations found was the sampling method being biased because people tended to share the survey with people who had similar characteristics. Therefore, the sample may represent specific population groups more distinctly. A second limitation is that the results obtained are not representative for the country as a whole. The collection, filtering, processing, and analysis of information and data was carried out by The Research and Project Management team at Profamilia in Bogotá, D.C. The research protocol was approved by Profamilia’s Research Ethics Committee (CEIP 14-2020 05) in the August 25, 2020 session.

The sociodemographic characteristics considered were age, gender, area of residence, vulnerability groups, level of education, ethnicity, marital status, department (region) of residence, type of health insurance, employment status, and household income. Perception of risk and health was measured by perceived susceptibility. To establish susceptibility, people were asked about their perception on their perceived likelihood of being infected with COVID-19, their mental and sexual, and reproductive health under current preventive measures imposed by the Colombian government.

Behavioral changes included the perceived effectiveness and adoption of preventive behaviors to avoid infection and subsequent transmission, based on three question categories: 1) hygiene practices; 2) travel suspension; 3) physical distancing. Willingness and ability to engage in mandatory isolation and physical distancing was measured using the following question: During the past 5 months, which of the following measures have you personally taken to protect yourself and others from COVID-19? This includes not going to work, school, or other public places, and avoiding public transportation or cabs.

Community engagement was measured by two categories of questions: actions taken and community engagement. For measures taken, the survey asked: 1) In your neighborhood, community, apartment complex, township, or district, have measures been taken or have campaigns been conducted to prevent the spread of COVID-19?; 2) What measures that you know of have been implemented to prevent the spread of COVID-19? and 3) What kind of measures would you like to see implemented in your community to keep you safe and reduce any risks? To measure engagement, we asked: How would you like to be involved in the community to support and cope with the pandemic? Immediate changes and expectations were measured by questions regarding: i) what people consider acceptable situations regarding education, work, and daily life in a COVID-19 future; ii) financial difficulties, loss of employment, and expectations about the vaccine and the so-called return to “normal”.

## Results

The results are presented in the following order:

1. Socio-demographic characteristics.
2. Changes in personal response and compliance with measures.
3. Changes in compliance with mandatory quarantine.
4. Changes in the three groups reacting to quarantine.
5. Changes in burden of care and employment at home.
6. Changes in perception of risk.
7. Changes in unmet sexual and reproductive health needs.
8. Changes in the mental health issues.
9. Changes in access to quality information.
10. Changes in citizen perception of government response.
11. Community engagement.
12. Expectations about children’s education, teleworking, and the possibility of a near-future without a vaccine or treatment.

### 1) Socio-demographic characteristics

17% of the people who responded to the survey had participated in the previous survey, Solidaridad I; in general, the survey was filled out in higher percentages by urban residents (95%), women (78%), people under 39 years of age (75%), single/never married people (54%), people with university studies (including technical/technological studies) (56%) and people with family incomes under five million pesos (73%). Figures 1-3 describe the socio-demographic characteristics of the people who responded the survey.

**Figure 1.**
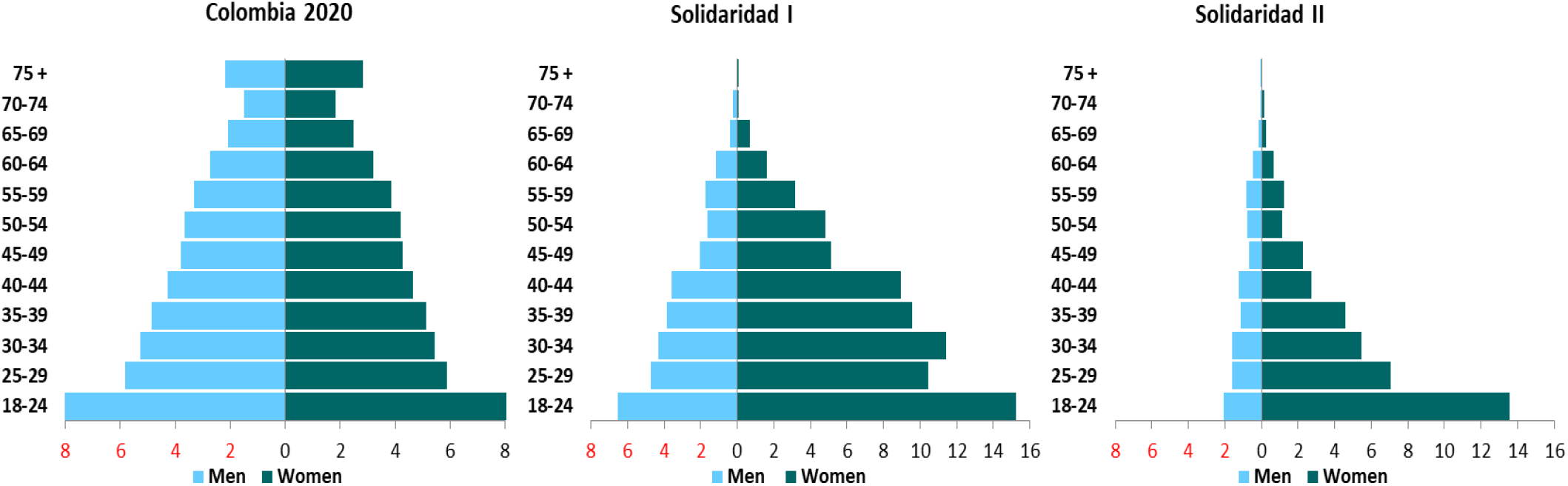
Population pyramids for respondents of Solidaridad Study I, II and at the overall national population according to DANE (National Department of Statistics and Census).

**Figure 2.**
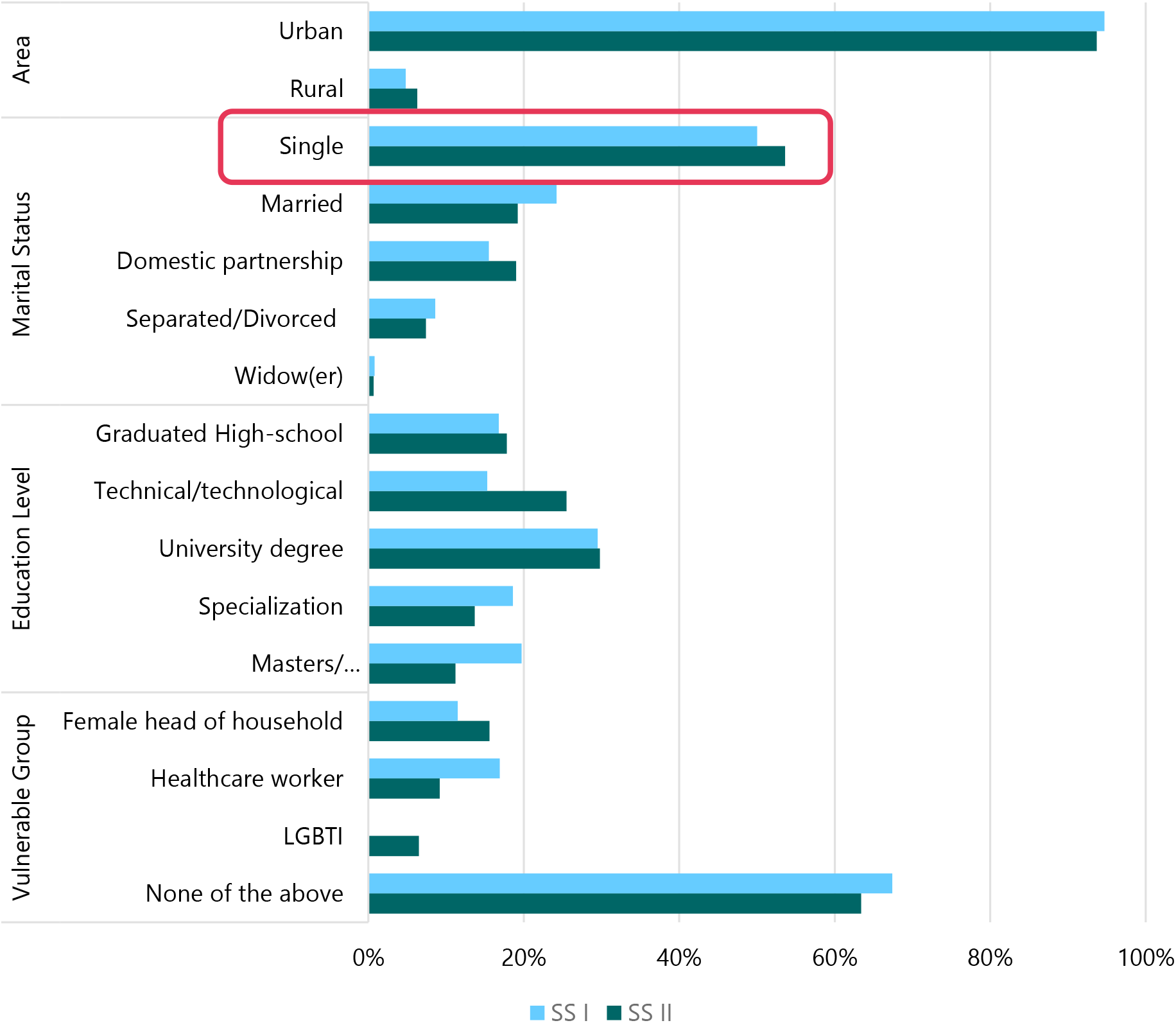
General characteristics of Respondents to Solidaridad Study II

**Figure 3.**
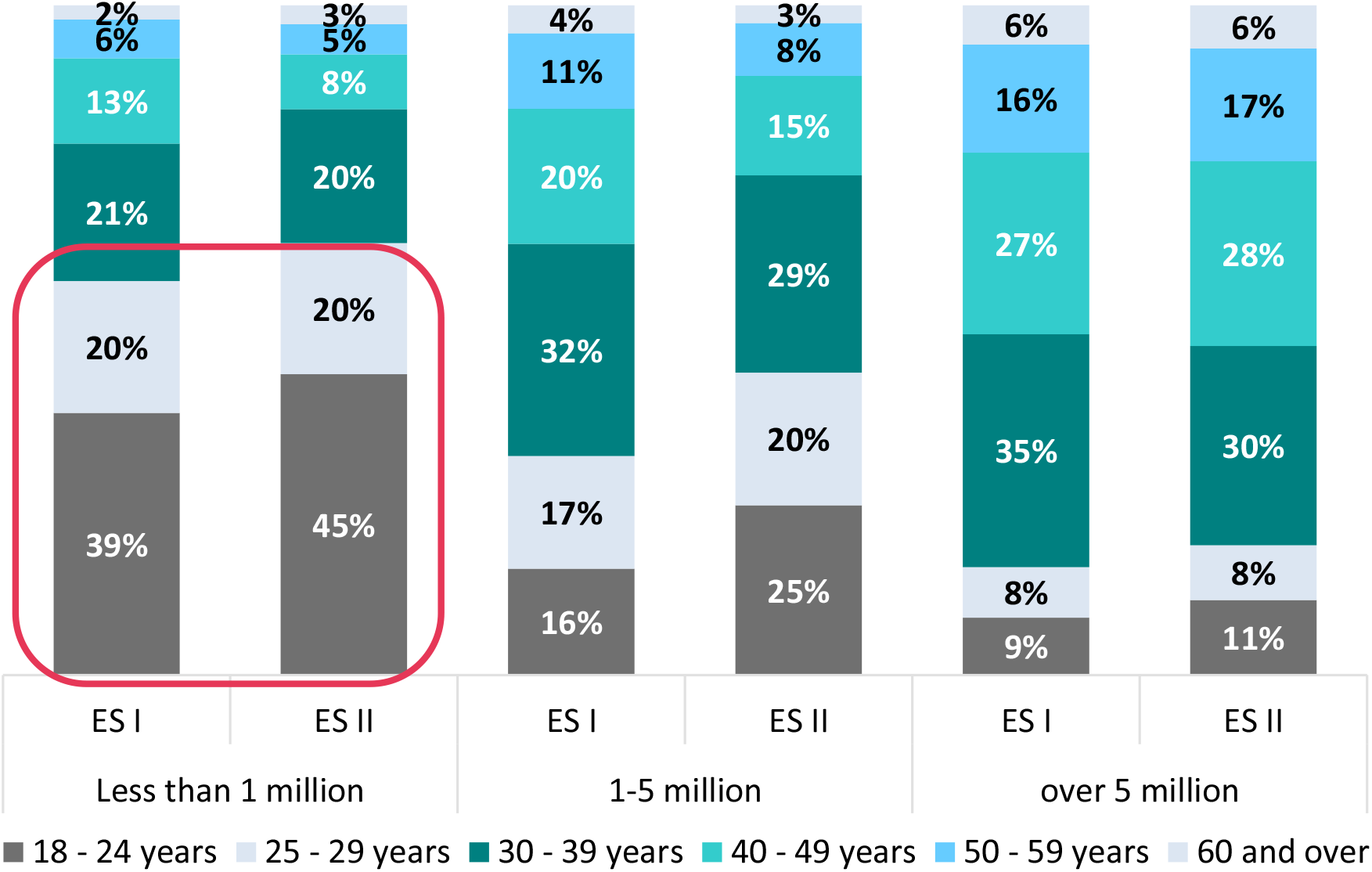
Income Level of Respondents’ Families by Age Group.

### 2) Changes in Personal Response and Compliance with Measures

An increase can be observed in the percentage of people who adopted measures to protect themselves and others against COVID-19 from the start of the quarantine when compared to the relaxation stage. The largest increases were reported in the use of face masks (28%), travel suspensions (20%) and going out to social events (11%). (Figure 4).

**Figure 4.**
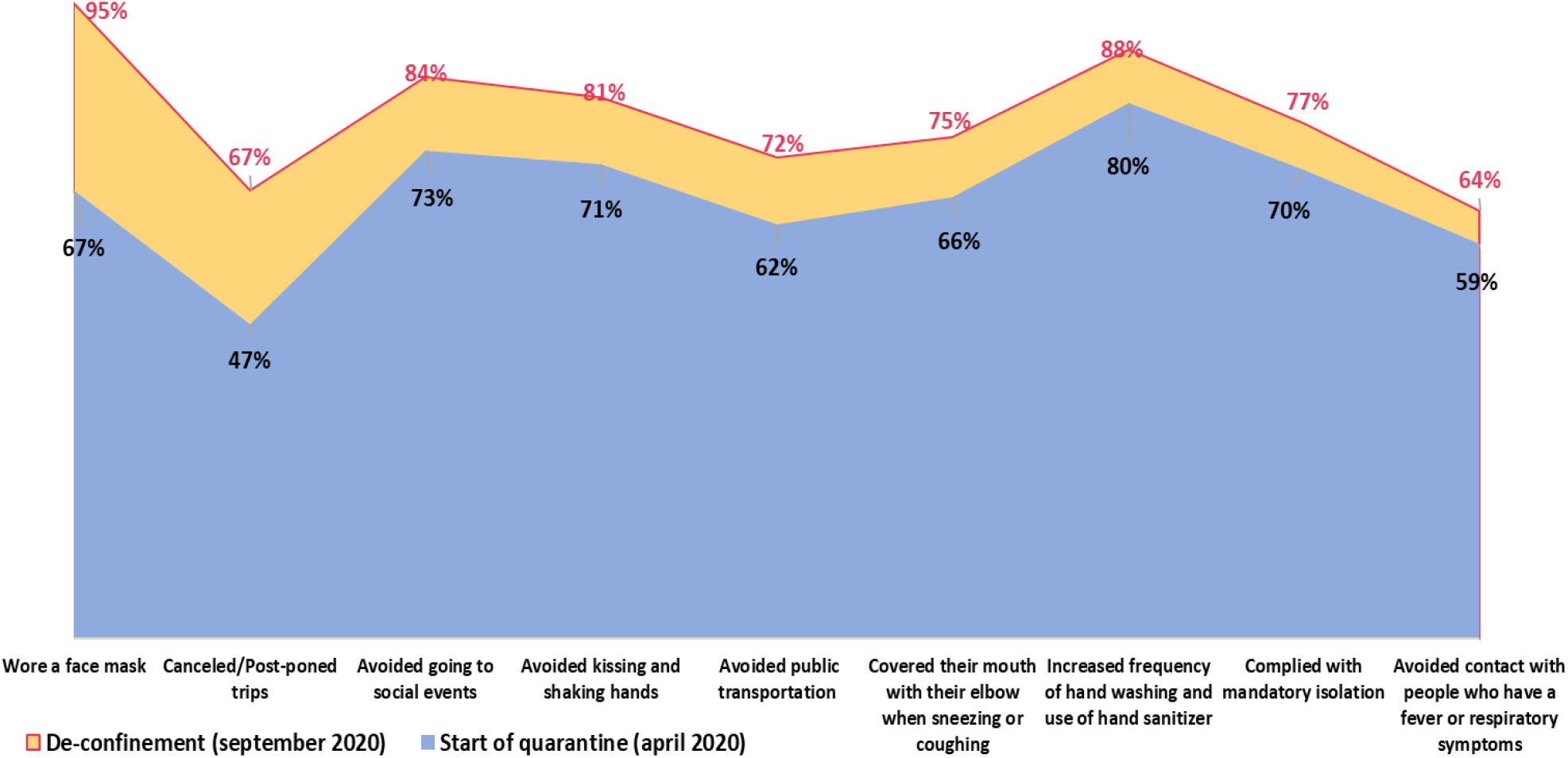
Changes in the Percentage of people who adopted measures to protect themselves and others from COVID-19 during the start of the quarantine and post-quarantine, Colombia 2020.

- People between 40 and 59 years of age adopted hygiene and self-care practices as well as preventive behaviors associated with physical distancing in greater percentages to prevent COVID-19.
- By gender, the most significant differences between the adopted behaviors were found in changing clothes upon returning home, which was 59% among men and 69% among women; in washing or disinfecting items taken home, 71% for men and 82% for women; and in increasing the frequency of cleaning at home/work, 56% among men and 66% among women.

### 3) Changes in Compliance with the Mandatory Quarantine

77% adopted the government’s mandatory preventive isolation; of these people, 42% started working from home. The percentage of people who began working from home is lower among women heads of household (26%) and victims of the armed conflict (28%), which reveals that conditions for informal workers and vulnerable populations is less flexible.

- People between 55 and 59 years of age (94%) and between 45 and 49 years of age (92%) adopted the mandatory isolation decreed by the government in higher percentages.
- 77% of women and men and 100% of people with non-normative genders adopted the mandatory self-isolation decreed by the government.
- 78% of people who do not work, 77% of people who have a paid job and 75% of those who work without pay adopted the mandatory isolation measure.
- 88% of people with a combined monthly family income of more than 5 million pesos and 75% of people with a combined family income of less than 3 million pesos went into mandatory quarantine.

9% of people have not complied with mandatory isolation.

- 75% left home to stock up on food
- 65% to go to work/university/college
- 41% to go to the bank
- 34% to buy medicines
- 33% to buy sanitizer, face masks
- 33% to attend a health facility
- 25% to walk their pet
- 24% due to anxiety and depression issues
- 23% to walk or exercise
- 22% to visit someone else’s home
- 15% to look for a job
- 12% to go to shopping malls or stores
- 10% to attend social gatherings
- 9% to help a person they are responsible for
- 8% to go to a notary

Of the women who did not comply with mandatory isolation, 70% left to go to work, 38% of men left their home because of anxiety and depression, and 35% went out to walk or exercise.

- 84% of the young people (18-24 years of age) went out to stock up on food, 43% due to anxiety and depression issues and 50% to visit someone else’s home.
- 86% of people between 30 and 49 years of age and 86% of those between 50 and 59 years of age left their homes to go to work.

The main worries raised due to the emergence of COVID-19 and the resulting isolation included

- 89% a family member getting infected.
- 87% the need for a medical emergency not receiving care.
- 86% how the pandemic could affect the poorest and most vulnerable people.
- 86% the future of the economy and economic recession.
- 83% people’s failure to comply with government measures.
- 81% uncertainty about the return to normal life.
- 79% availability of a vaccine or treatment soon.
- 79% availability of a vaccine or treatment in Colombia
- 74% access to a vaccine or treatment once either arrives in Colombia.
- 72% lack of food, medicine and medical supplies.
- 67% family members living alone.
- 67% not being able to pay rent or public services.
- 66% loss of employment, income and savings.
- 65% falling into anxiety and depression during isolation.
- 45% being alone and unable to take care of themselves.
- 37% educational future of their children.
- 24% not having a computer or internet access.
- 23% domestic violence.
- 23% an impact on children’s behavior.
- 20% an impact on social life.

The percentage of people who expressed some concern due to COVID-19 increased from at the start of the quarantine and during the relaxation stage according to the surveys. (Figure 5).

**Figure 5.**
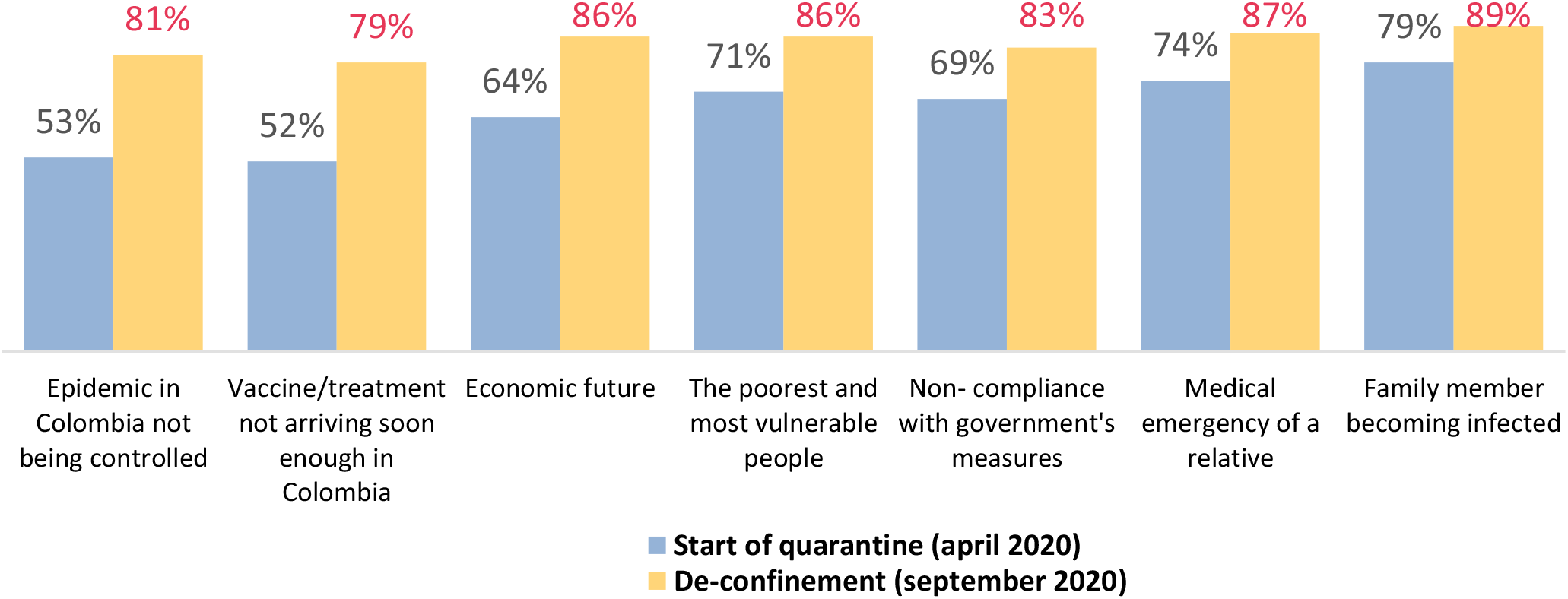
Changes in the Concerns Raised by the Emergence of COVID-19 and Mandatory Isolation, Colombia 2020.

- 28% increased uncertainty regarding the return to normal life as a result of the epidemic not being under control in Colombia.
- 27% were worried the vaccine or treatment would not arrive soon enough in Colombia.
- 22% were worried about their financial future.

### 4) Changes in the Three Groups Reacting to Quarantine Measures

A preliminary examination of the results using the *k-means* technique helped identify three groups that are reacting in different ways to the pandemic, changing their behavior, and facing physical distancing measures (4):

- Those who accept it (33%)
- Those who resist the situation (30%)
- Those who suffer because of it (37%).

Some characteristics of each of the three groups are highlighted below.

#### Group 1. Those Who Resist

The group of people who resist is made up by 29% men; 52% are in the 30-39-year age group; 29% are married, 37% have family incomes between $1 to $3 million pesos per month and 62% have not had chronic illnesses. In their response to COVID-19, this group is characterized by the following behaviours:

- 20% adopted four or fewer physical distancing measures. In the lowest percentages:
- 90% adopted hygiene measures.
- 86% avoided going out to social events and 74% avoided using public transportation.
- 73% avoided contact with people who have fever or respiratory symptoms.
- 45% avoided contact with people who had traveled in five months leading up to the survey. 99% have experienced at least one mental health event during the pandemic. 85% have experienced four or more mental health events during the pandemic.

#### Group 2. Those Who Suffer

In the group of people suffering from the pandemic 79% are women, 68% are under 30 years of age, 69% are single, 36% have an average family income of less than $1 million pesos per month, 60% have had some chronic disease. In their response to COVID-19, this group is characterized by the following behaviours:

- 86% adopted more than three physical distancing measures.
- 88% have had at least one mental health event during the pandemic.
- 95% felt tired for no apparent reason.
- Over 95% are very concerned about:
  - Someone in my family getting COVID-19
  - Themselves or someone in my family having an emergency and not receiving care
  - Their future finances and economic recession
- One in four of the poorest and most vulnerable people have suffered from psychological violence.

#### Group 3. Those Who Accept

In the group of people who adapt to the situation, 80% are women, 41% are under 30 years old, 45% are in the 30-49 years of age group, 44% are married or living in domestic partnerships, and 67% have not had chronic illnesses. In their response to Covid-19 this group is characterized by the following:

- 87% adopted more than three social distancing measures
- 97% have taken at least one distancing measure.
- In higher percentages:
  - 73% feel that by complying with mandatory social isolation, they are contributing to stopping the spread of COVID-19.
  - 67% Watches movies/series.
  - 65% Can spend time with their family.
  - 64% Avoid using public transportation and exposure.
  - 55% Communicate daily with family members, friends (office colleagues).
  - 50% Use social media to stay informed and connected.
  - 49% Stopped listening or reading the news.

Compared to the beginning of the quarantine (five months later), the percentage of people suffering from the pandemic increased by 11%, the number of people who resist it decreased by 4%, and the percentage of people who accept the pandemic decreased by 7%. (Figure 6)

**Figure 6.**
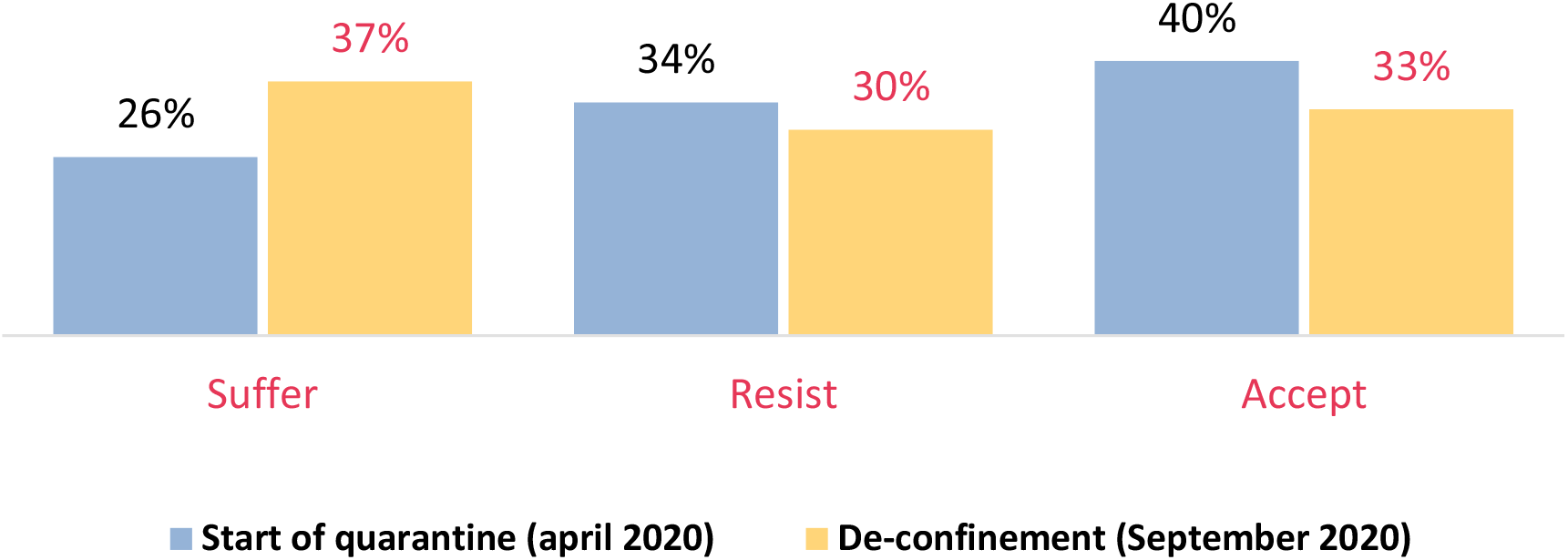
Three Groups of Response to the Pandemic and Adopted Measures, Colombia 2020.

### 5) Changes in Burden of Care and Employment at Home

68% are responsible for the care of a member in their household. 22% takes care of a child younger than 5 years of age.

- 37% are in charge of the care of children between 5 and 17 years of age.
- 43% are in charge of the care of people between 18 and 64 years of age.
- 24% are in charge of the care of an elderly person between 65 and 84 years of age.
- 5% are responsible for the care of people over 85 years of age.
- 5% are responsible for the care of a disabled person.

68% of women and 68% of men are responsible for the care of a person in their household. 77% adhered to the mandatory isolation decreed by the government; of the people who left their homes during quarantine, 75% left to buy food, 65% to go to work/school/university and 41% to go to the bank.

16% of the women are heads of household and among them, 95% are responsible for the care of someone at home.

- Among women heads of household who are responsible for someone at home, 41% have an average family income of less than one million pesos per month; 26% started teleworking, 43% are unemployed.
- 74% adhered to the mandatory isolation; 80% left their homes to go to work/school/university, 75% to buy food, and 40% to go to the bank.
- 51% are responsible for the care of children under 5 years old, 76% for the care of children between 5 and 17 years old, 67% for the care of people between 18 and 64 years old, 31% for the care of people between 64 and 84 years old, 10% for the care of people over 85 years old, and 8% for the care of disabled people.

26% reports an income of over 3 million pesos per month.

- 40% of men and 22% of women report an average monthly family income of more than 3 million pesos.
- The percentage of income over 3 million pesos increases with age.

### 6) Changes in Perception of Risk

94% see COVID-19 as a serious or very serious health problem; 74% think that COVID-19 can have serious health consequences for the whole population without distinction; 22% think that COVID-19 can have serious health consequences for some risk groups such as adults over 65 years of age.

- People feel that it is very likely or likely to become infected with COVID-19 by having contact with someone who is infected (87%); by not wearing a face mask (84%); by using public transportation (82%); by being within 2 meters of someone who has COVID-19 and coughs or sneezes (82%); by visiting crowded places such as marketplaces, malls, supermarkets (82%) and by having contact with contaminated surfaces (81%). (Figure 7).

**Figure 7.**
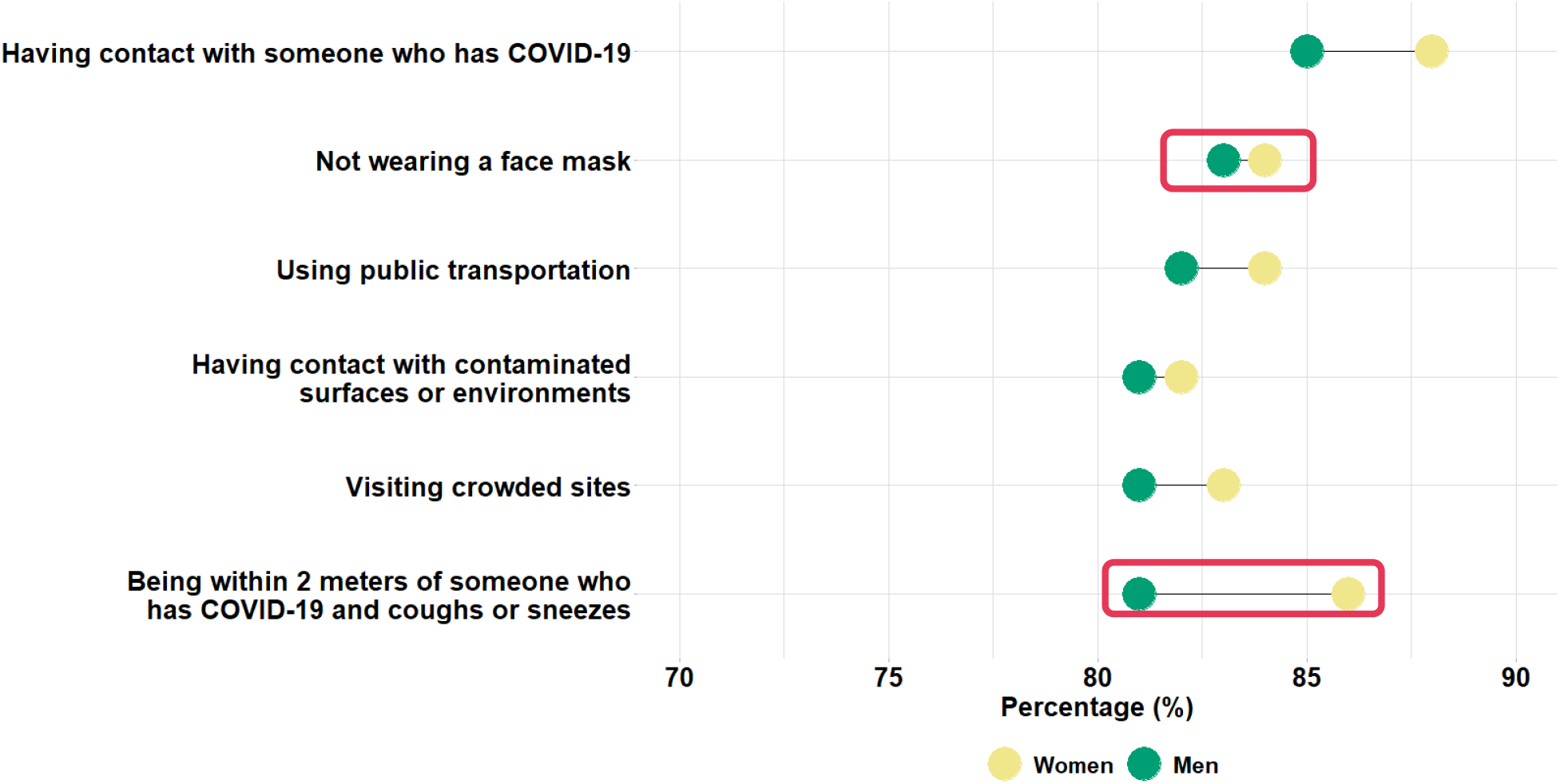
Percentage of People Who Consider It Very Likely or Likely to Become Infected with COVID-19 for Being Exposed to the Following Situations (by gender), Colombia 2020.
- The percentage of people who have not been tested for COVID-19 decreased by 38% compared with the beginning of quarantine, while the percentage of people who think COVID-19 is transmitted by being within two meters of someone who is infected and coughs and sneezes increased by 15% (Figure 8).

**Figure 8.**
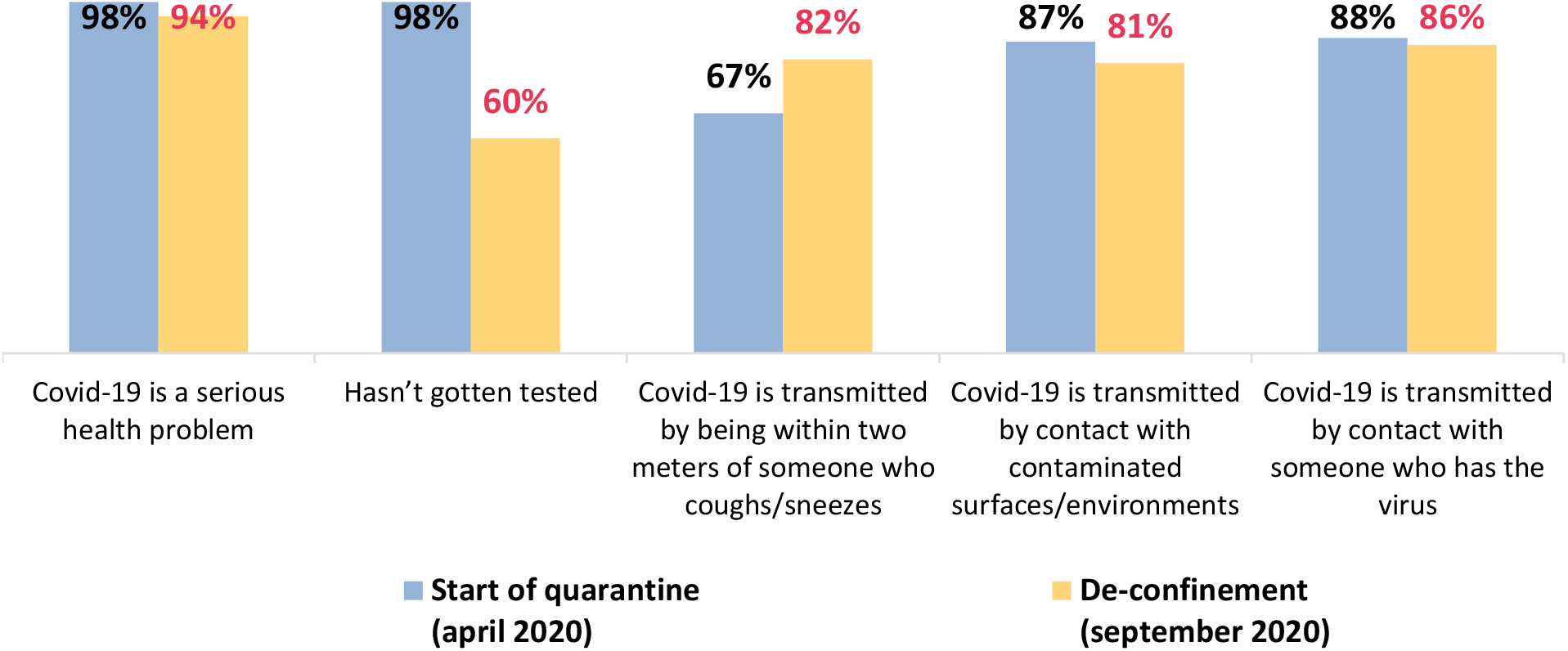
Changes in the Percentage of People Who Consider It Very Likely or Likely to Become Infected with COVID-19 after Being Exposed to the Following Situations, Colombia 2020.
- Regarding the perception of mental health: 43% said that their mental health condition has been affected in the last three months of 2019 (including depression, anxiety, or nervousness).
- 40% of people say that they or someone in their family has been tested for COVID-19, while, 60% has not been tested, and 17 people chose not to answer the question.
- 17% were tested and received the results in less than a week; 15% were tested and received the results one to two weeks later; and 8% were tested and received the results one month or more after being tested.
- 44% mentioned some of the diseases considered in the survey; among the most frequent we found:
  - 17% Mental illness (depression, anxiety, schizophrenia, loss of sleep).
  - 16% Ear, nose or throat disorders (e.g., rhinitis, deafness, tinnitus)
  - 7% chronic diseases.
  - 7% cardiovascular diseases.
  - 5% intestinal problems.
  - 5% respiratory diseases.

### 7) Changes in Unmet Sexual and Reproductive Health Needs

Regarding Sexual and Reproductive Health (SRH) attention needs in the COVID-19 context presented as follows:

- 39% of people have had some need for sexual and reproductive health (SRH) care.
- Among young people (18 to 29 years of age) (51%), people with a family income of less than one million pesos per month (48%), people from the LGBTI community (48%) and women (43%) had the highest percentages related to sexual and reproductive health service needs. Of the people who had some need for sexual and reproductive health care, a little more than half did not receive the required treatment(s) (53.4%).

Amongst women, the main sexual and reproductive health needs are:

- 27% gynecological visits.
- 20% access to contraceptives (condoms, IUD, pills, injections, implants, etc.)
- 16% contraceptive visits.

53% of the women who had some SRH-related need did not get proper treatment. Among the main causes for not getting appropriate treatment, we found that:

- 30% did not get treatment because they preferred not to leave home until the end of the quarantine.
- 29% did not get treatment because health care providers suspended their health care services as a result of the quarantine.
- 28% did not get treatment because they did not know whether their health care service was available (lack of information).
- 27% did not get treatment because they did not have the money to pay for the service and/or product.
- 21% did not get treatment because their health care provider has not authorized it or has delayed it.

Amongst men, the main sexual and reproductive health needs are:

- 9% Access to contraceptives (condoms, IUD, pills, injections, implants, etc.)
- 9% Urological visits.
- 7% Diagnostic tests for Sexually Transmitted Diseases.
- 6% Medical consults regarding the use of Contraceptives.

57% of the men who had some SRH-related need did not get proper treatment; among the main causes for the inadequacy of treatment, we found that:

- 39% did not get treatment because they opted to remain at home for the duration of the quarantine.
- 35% did not get treatment because health care providers suspended their health care services as a result of the quarantine.
- 33% did not get treatment because they did not know whether their health care service was available (lack of information).
- 23% did not get treatment because their health care provider has not authorized it or has delayed it.

Among non-normative gender individuals (11 people), 6 people claimed to have at least one sexual and reproductive health need; only 3 were able to get treatment(s). Among the reproductive health needs were: 4 cases of people needing gynecological visits; 3 cases of people needing access to contraceptives; 3 cases in which individuals needed STD screenings; 2 cases in which respondents needed contraceptive visits and 1 case in which the person needed endocrinological consults (hormonization, hormone replacement therapy). Some of the reasons why individual needs have not been met are as follows: 2 people did not access the services because they did could not afford the service; 2 people preferred not to leave home until the end of the quarantine; in 1 case, they did not know whether the service was available (lack of information); in 1 case, the service was not attainted because the service was not available in their municipality and finally, in 1 case, the service hours during the quarantine did not adapt to the study and work schedules of the respondent.

Sexual and reproductive health needs have increased since the beginning of the quarantine, mainly those in gynecology services (12%), access to contraceptives (12%) and contraceptive visits (11%) (Figure 9).

**Figure 9.**
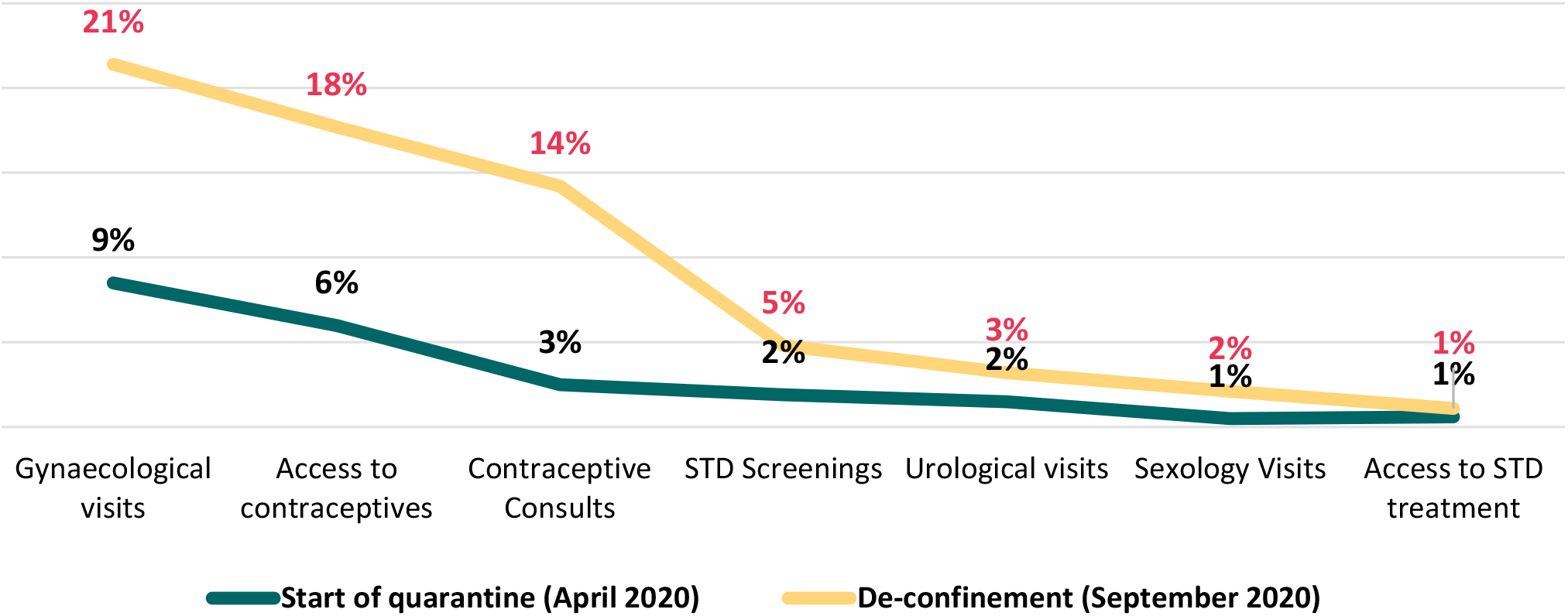
Changes in the Percentage of People who Had Sexual and Reproductive Health Needs During Quarantine, Colombia 2020

6% of people have had to change their contraceptive practices as a result of the pandemic. 7% were using contraceptives before the pandemic and stopped using them during the pandemic. 38% were not using contraceptives at all.

- Among women, 6% changed their contraceptive practices due to the pandemic.
- 8% stopped using them.
- 36% were not using any contraceptives.
- Among men, 3% changed their contraceptive practices.
- 2% stopped using contraceptives.
- 42% were not using any contraceptives.
- 7% of people with a monthly income of less than one million pesos had to change their contraceptive practices and made up the highest percentage among income groups.
- 9% of people with monthly incomes of less than one million pesos had to stop using contraceptives and represented the highest percentage among income groups.
- 42% of the people with monthly incomes over five million pesos were not using contraceptives and made up the highest percentage among income groups.

25% of people claimed to have decreased the frequency of their sexual activity during the pandemic, 32% claimed that their sexual activity did not change at all, and 17% stopped having sexual intercourse.

- One in three women said their sex life has not changed.
- One in three men stated that the frequency in which they engaged in sexual activity has decreased.
- 27% of young people (18 to 29 years of age) said that their sexual activity frequency has decreased since the pandemic started.
- 41% of people aged 30-49 said their lives have not changed.
- 35% of people over 49 years of age said that they are not sexually active.
- One in four single people decreased their sexual activity during the pandemic.
- One in four single people stopped having sex.
- 54% of married people said their sex lives have not changed.
- Half of those who were in a relationship said that their sexual activity had not changed.

### 8) Changes in Mental Health Issues

92% have experienced the impact of some form of mental health condition in the last month of quarantine. Figure 10 shows the percentage distribution of the main mental health conditions by sex:

**Figure 10.**
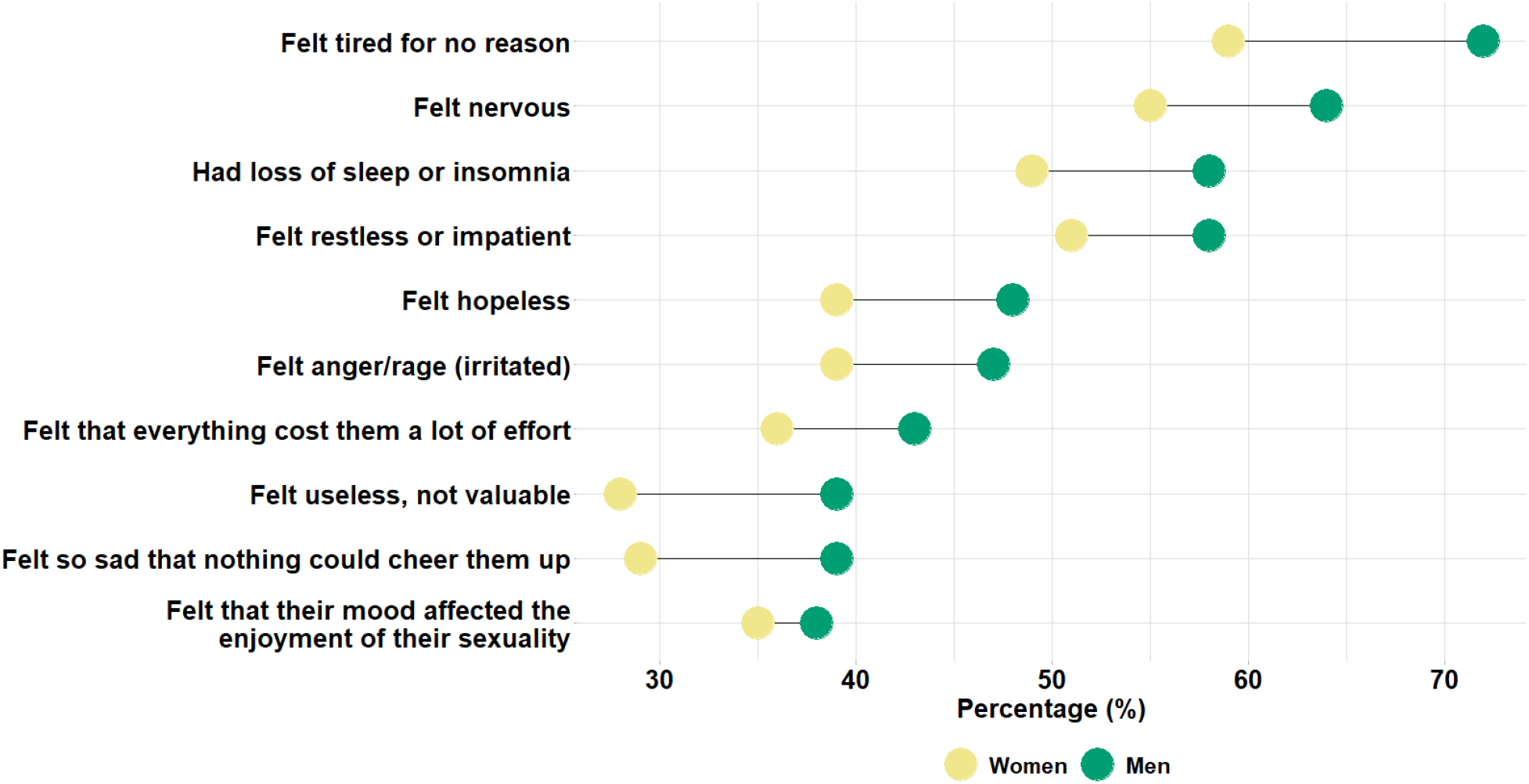
Percentage of People Who Have Felt Any of the Following Mental Health Conditions in the Last Month of Quarantine by Gender, Colombia 2020.

- 70% felt tired for no apparent reason
- 62% felt nervous
- 57% felt restless or impatient
- 56% suffered from sleep deprivation or insomnia
- 46% felt hopelessness
- 46% felt anger/rage (irritated)
- 42% felt that everything takes a lot of work
- 38% felt that their mood affected their sexual enjoyment
- 37% felt so sad that nothing could cheer them up
- 37% felt useless, worthless

Figure 11 highlights the most frequent types of effects on mental health experienced by respondents during the last month of the quarantine and categorizes results by age group:

**Figure 11.**
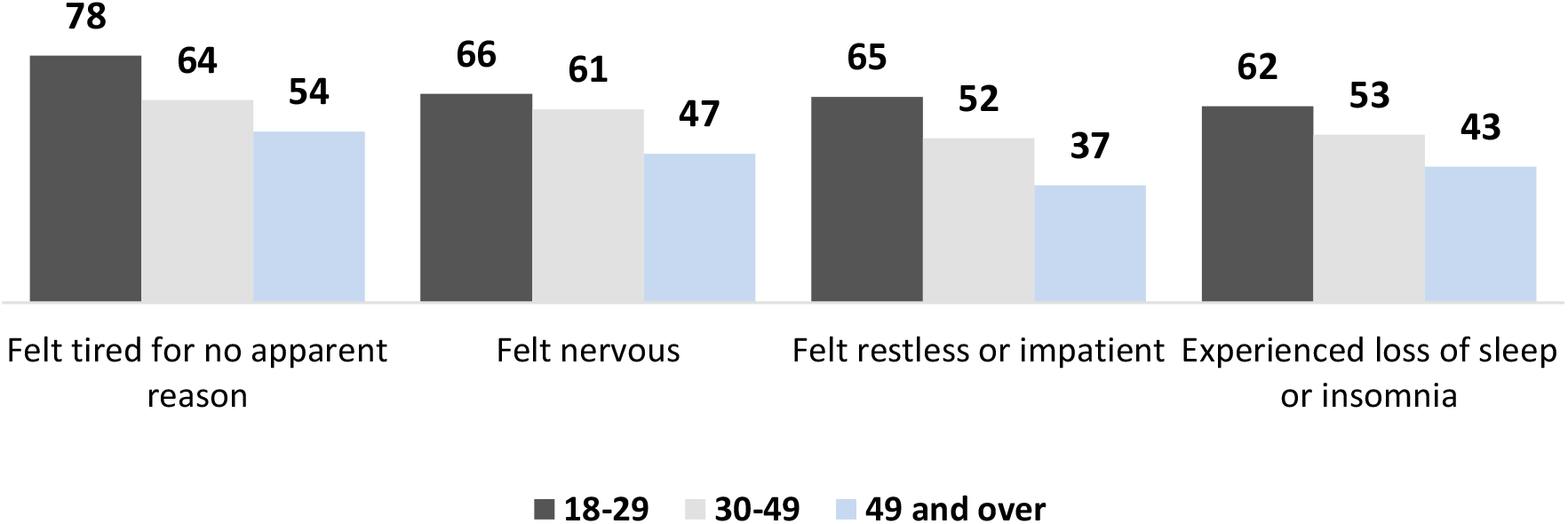
Percentage of People Who Have Experienced Mental Health Issues during the Last Month of Quarantine by Age Group, Colombia 2020.

- The 18-29-year-old group has often felt tired for no evident reason (78%), nervous (66%), restless (65%) and experienced loss of sleep (62%), compared to the 49+ age group who felt less tired, nervous, restless or did not experience loss of sleep in the same frequency.
- Among the 49+ age group, feeling tired (54%) was more common during the last month of quarantine and perhaps was the greatest reported mental health issue.

In general, mental health-related issues increased starting at the beginning of the quarantine and during its last month (Figure 12). The study found that there was a/an:

**Figure 12.**
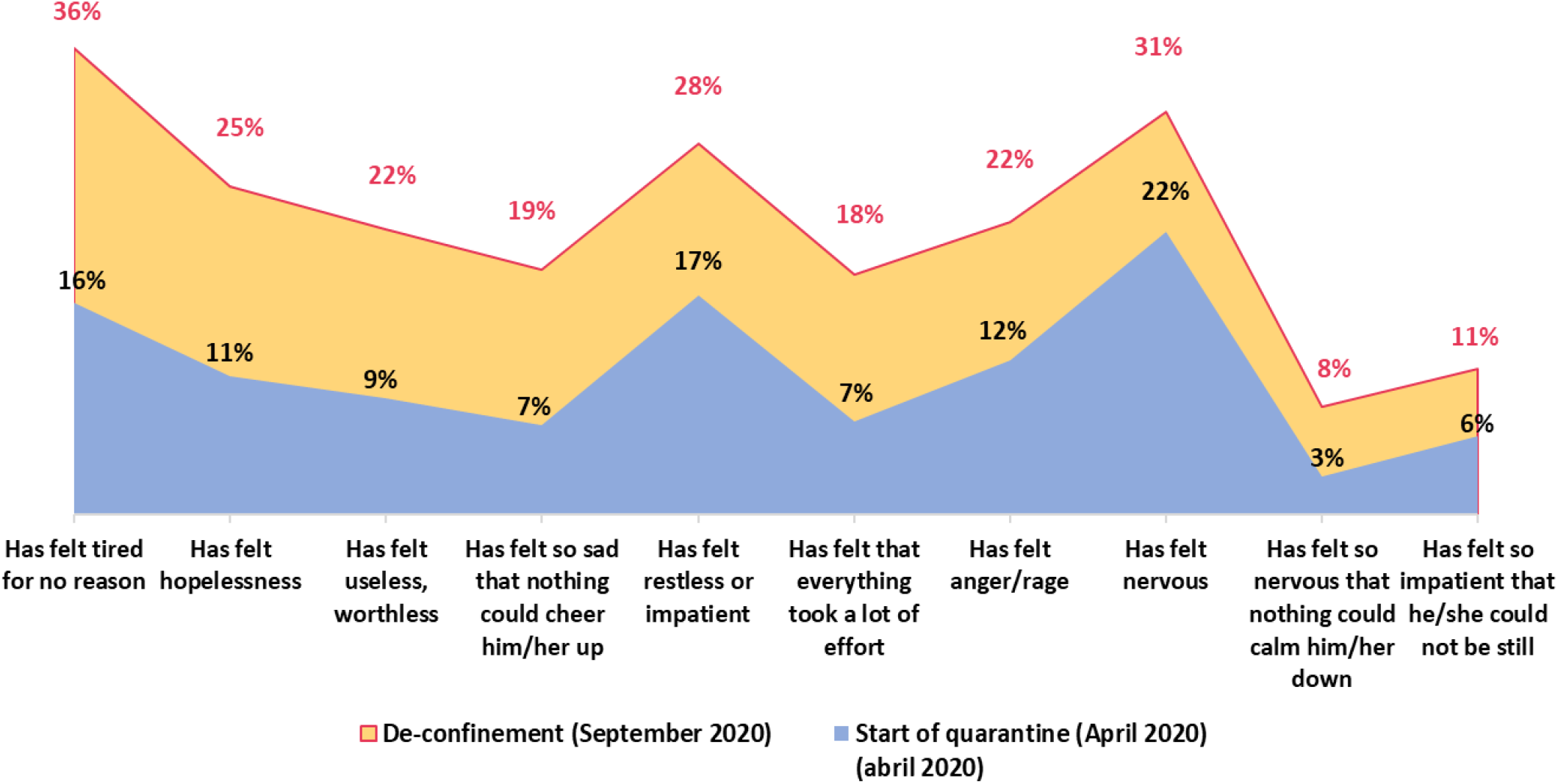
Changes in the Percentage of People Who Experienced Some Mental Health Issue at the Beginning and during the first and the Last Month of six-month Quarantine, Colombia 2020.

**Figure 13.**
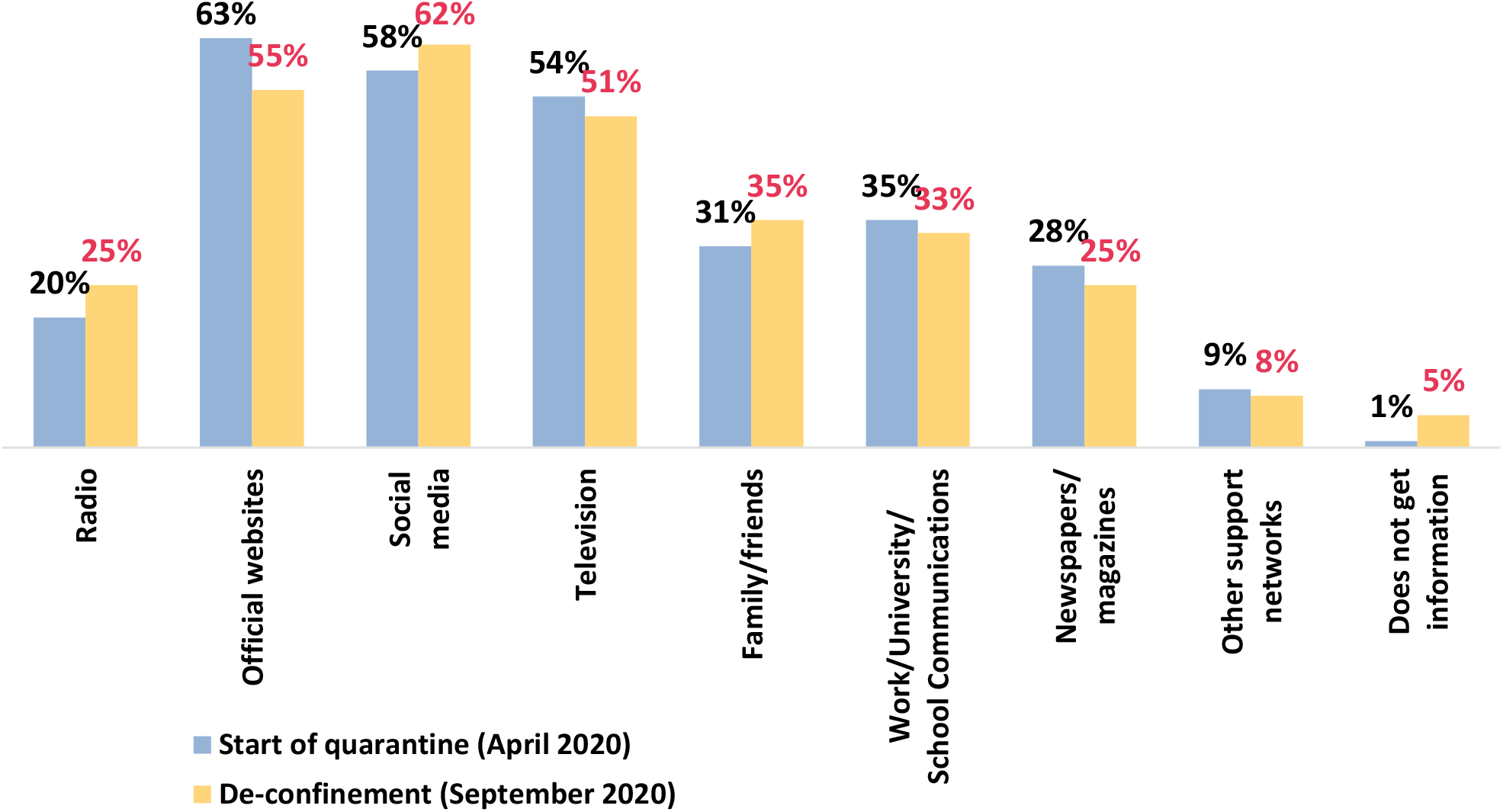
Changes in Information Sources

- 20% increase in feeling always or often tired for no reason.
- 14% increase in feeling always or often hopeless.
- 13% increase in feeling always or often useless or worthless.
- 12% increase in feeling always or often so sad that nothing could cheer the respondent up.
- 11% increase in feeling always or often restless or impatient.
- 11% increase in feeling always or often that everything cost them a lot of effort.

### 9) Changes in Access to Quality Information

People have received information about COVID-19, during quarantine, through the following sources:

- 55% Official websites
- 51% television
- 35% family and friends
- 33% statements from work, school or university
- 25% newspaper/magazines (printed or digital)
- 25% radio
- 9% other official websites
- 8% other support networks

No significant differences were found regarding respondent media preferences by gender; the three most frequently used media were the similar across groups. The following characteristics were found:

- Access to information about COVID-19 through social media is higher among young people under 25 years of age (70%) and decreases with age (45% in the group between 55 and 59 years of age); however, for people over 59 years of age, it is 61%.
- Access through official websites is higher among people aged 35-49 and lower among people aged 60 and over.
- Access to COVID-19 information through television increases with age (from 50% among those under 25 years of age to 66% among those over 59 years of age).
- People who did not adhere to mandatory isolation have the lowest percentages of access to COVID-19 information through any media source. Among the most used media sources, access through social networks reaches 64% for those who went into mandatory isolation, but drops to 55% for those who did adhere to isolation measures; access through official websites drops from 61% to 43%, and access through television from 53% to 43%.
- 5% do not receive any information about COVID-19.

### 10) Changes in Citizen Perception of Government Response

In Colombia:

- 31% strongly agree or agree that the national government’s response to control COVID-19 was clear and consistent.
- 32% strongly agree or agree that the national government acted in a timely manner to control the spread of COVID-19.
- 39% strongly agree or agree that the local government response to control COVID-19 was clear and consistent.
- 47% strongly agree or agree that the local government response to control COVID-19 was clear and consistent.
- Men have, in higher percentages, a favorable perception (strongly agree or agree) regarding the clarity of response and the timeliness of national and local government action.
- Among older people (49 years and older) there are higher levels of approval regarding response and action of the national and local governments.
- Antioquia and Bogotá have the highest levels of approval in terms of clarity of response (43% and 42% respectively) and the timeliness of local government action (51% and 56% respectively).
- Above 80% of the people who had a favorable perception regarding response and actions of their local government to control the spread of COVID-19 adhered to the mandatory isolation decreed by the government.

The favorable perception that people had at the beginning of the quarantine regarding the response and actions of the national and local governments to control COVID-19 decreased through the course of the quarantine (Figure 14).

**Figure 14.**
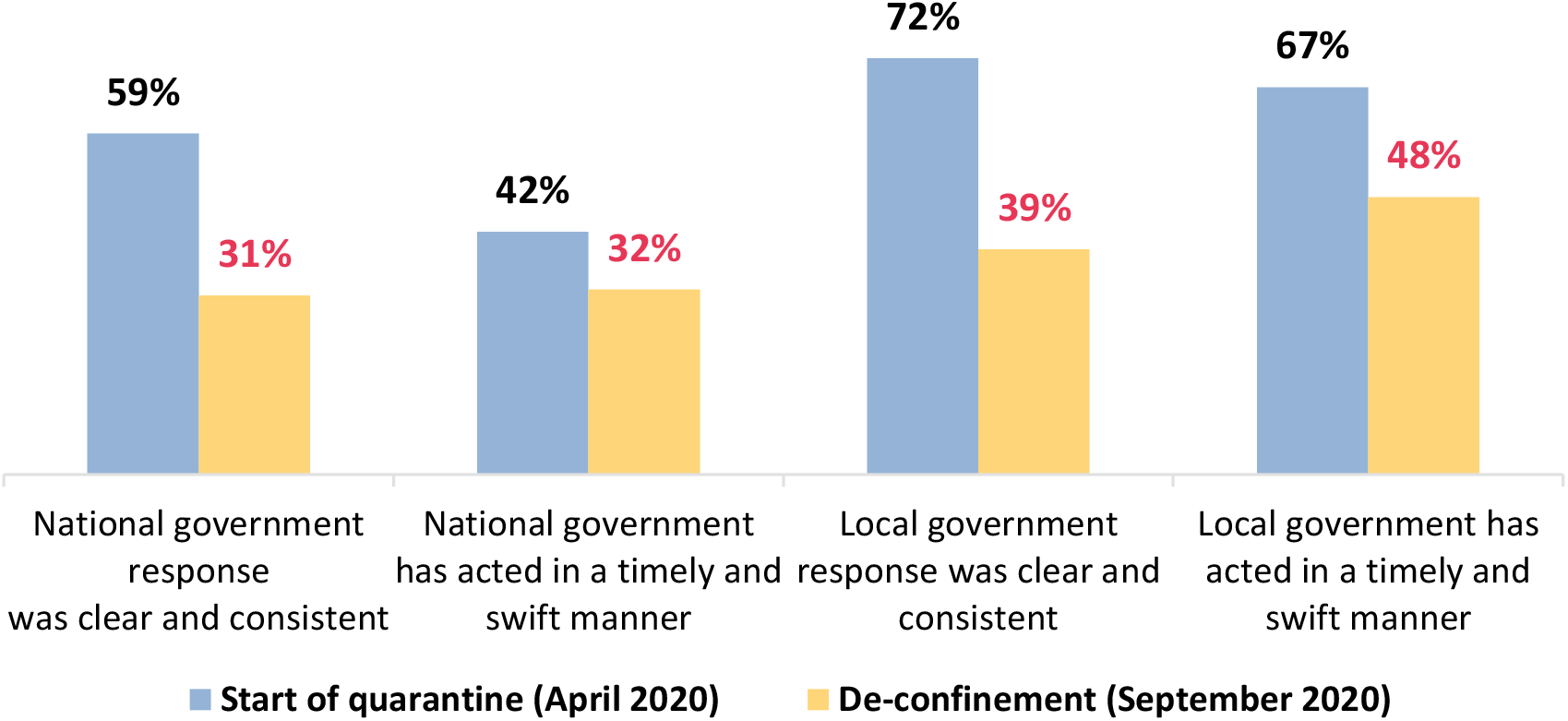
**Changes in the** Perception of National and Local Government Response to Control COVID-19 From the Start of the Quarantine and During the Post-Quarantine, Colombia 2020.

**Figure 15.**
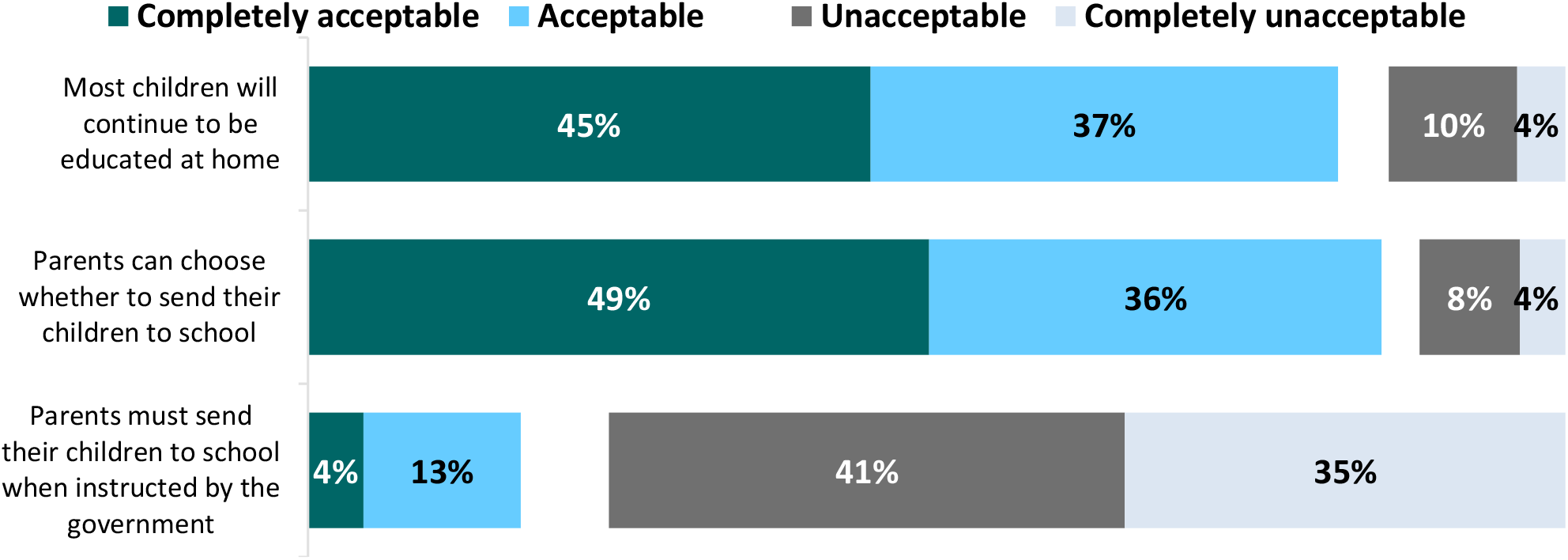
Acceptable Situations Related to Children’s Education, Colombia 2020.

**Figure 16.**
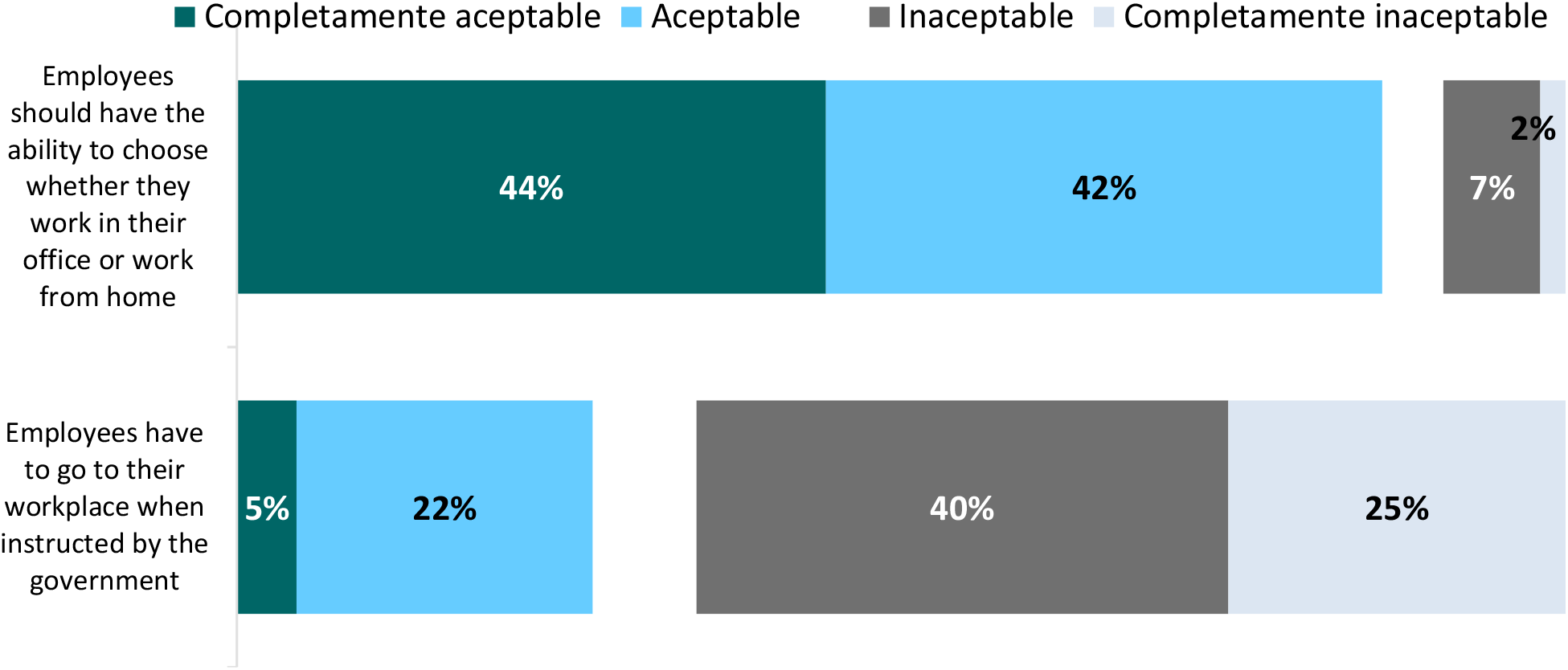
Acceptability of Work-related Situations. Colombia 2020

**Figure 17.**
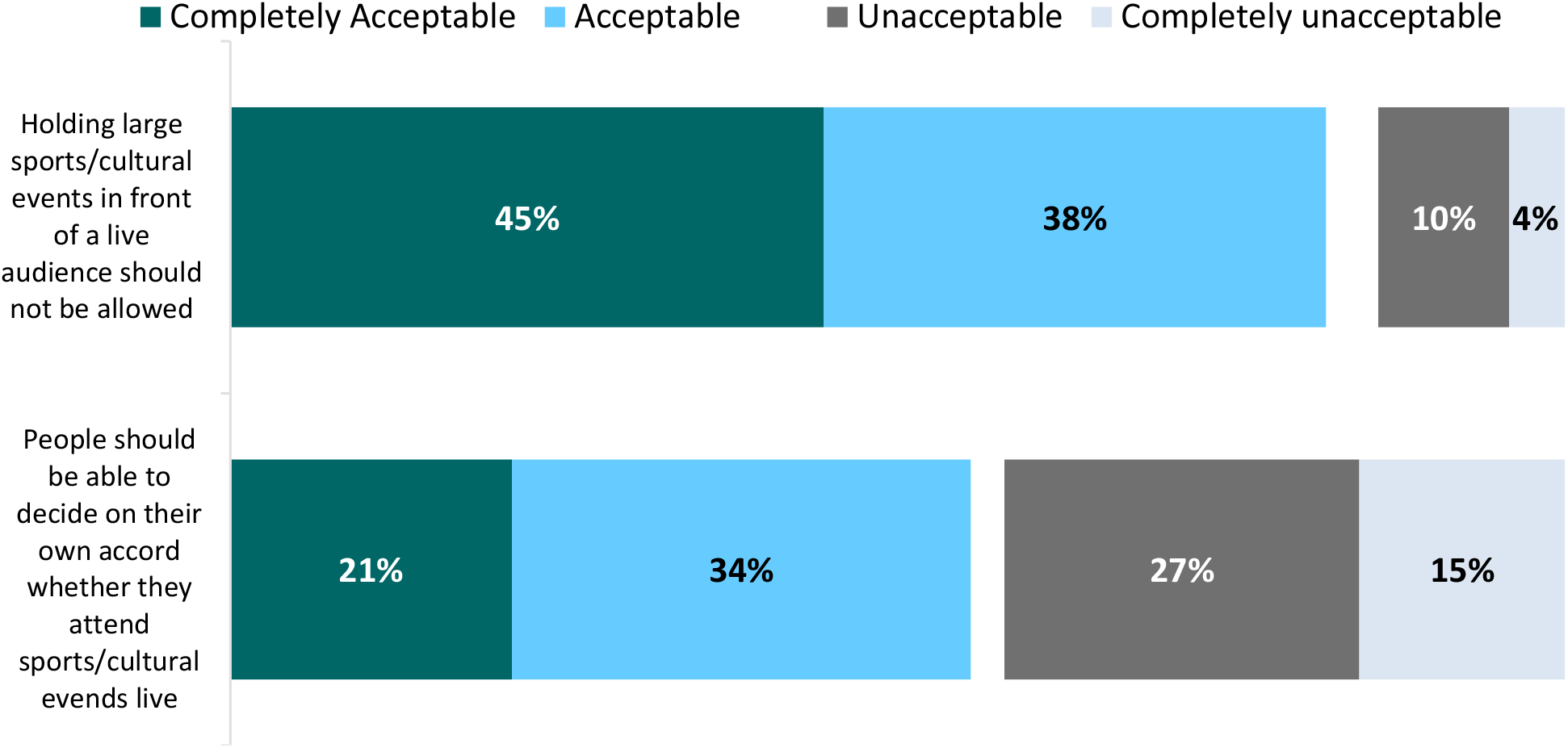
Acceptability of Situations Related to Daily Life, Colombia 2020.

**Figure 18.**
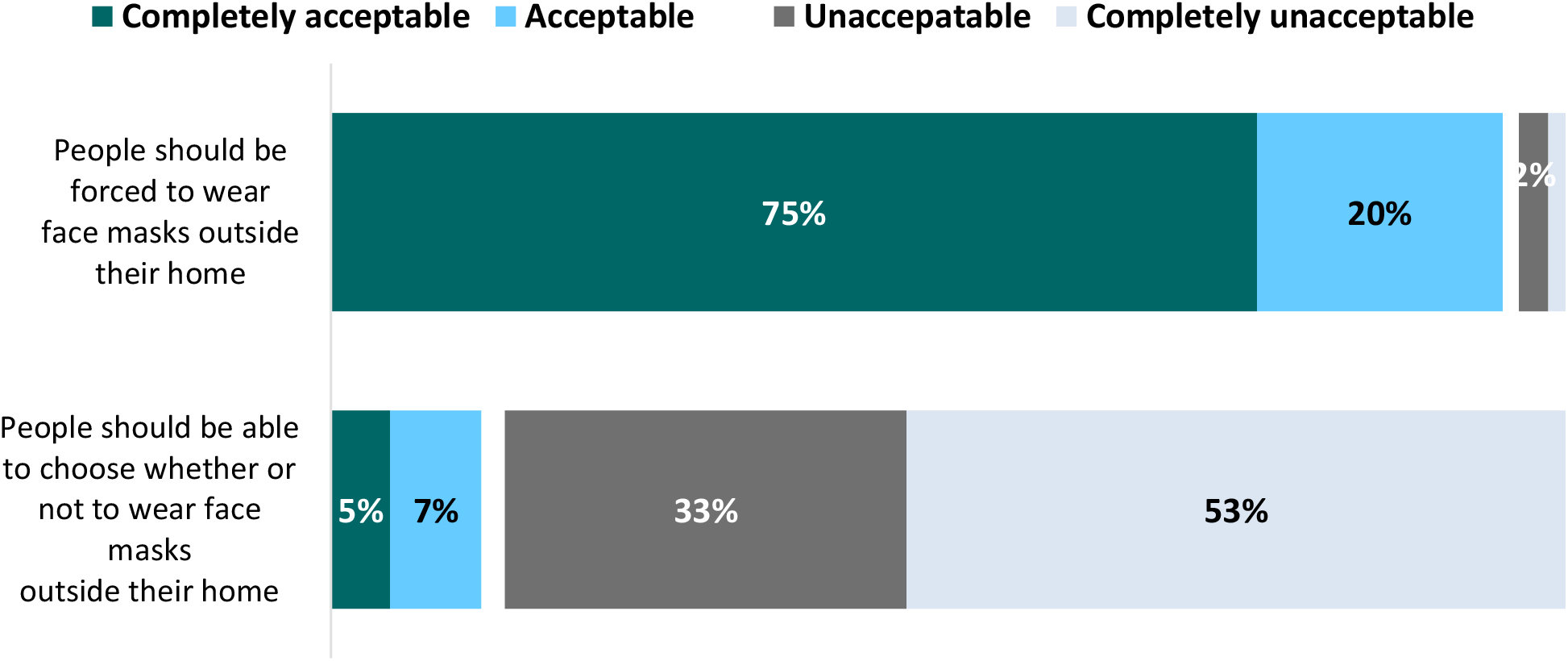
Acceptability of Situations Related to the Use of Face Masks, Colombia 2020.

**Figure 19.**
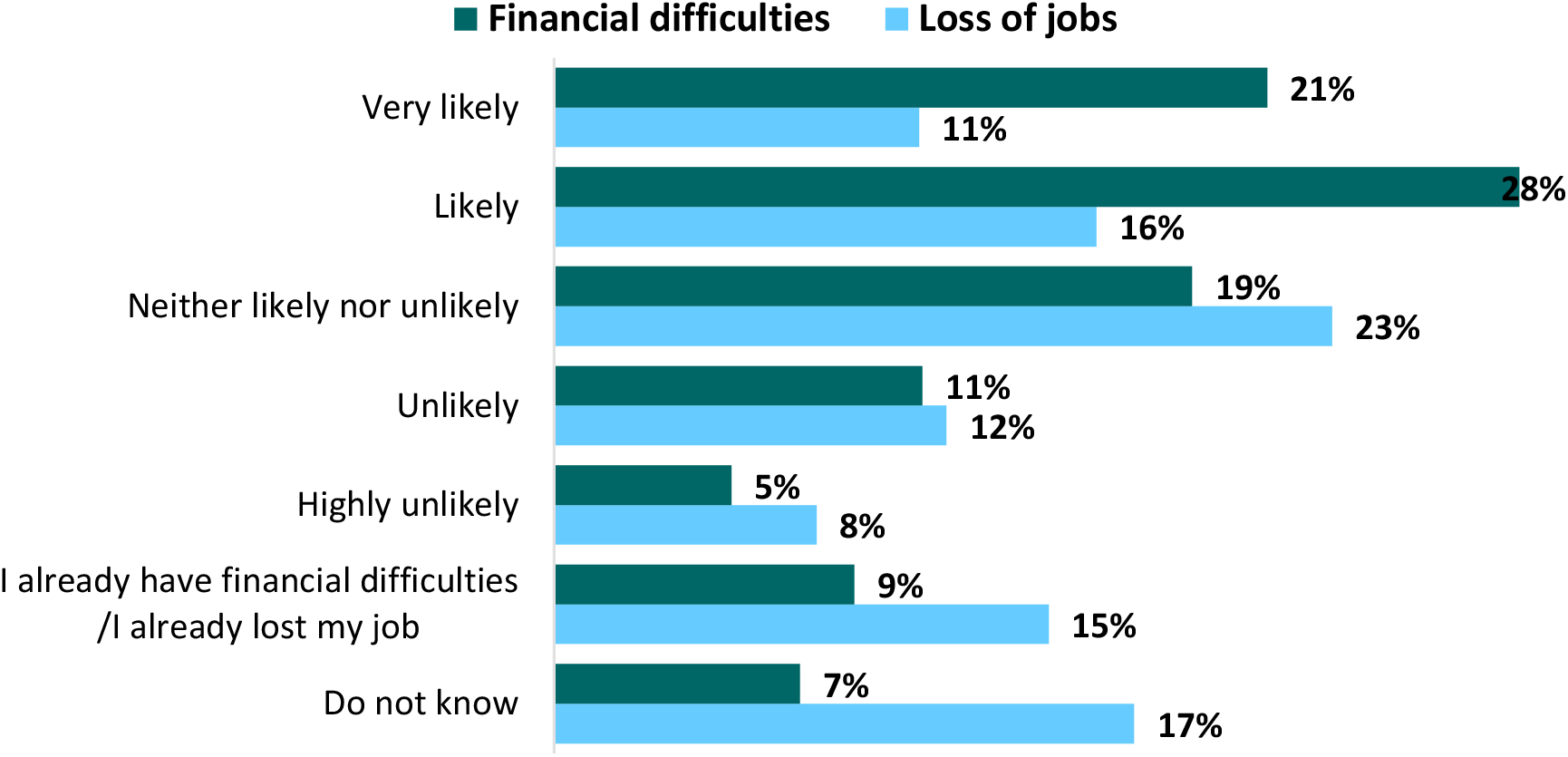
Probability of Facing Financial Difficulties and Job-Loss as a Result of COVID-19, Colombia 2020.

**Figure 20.**
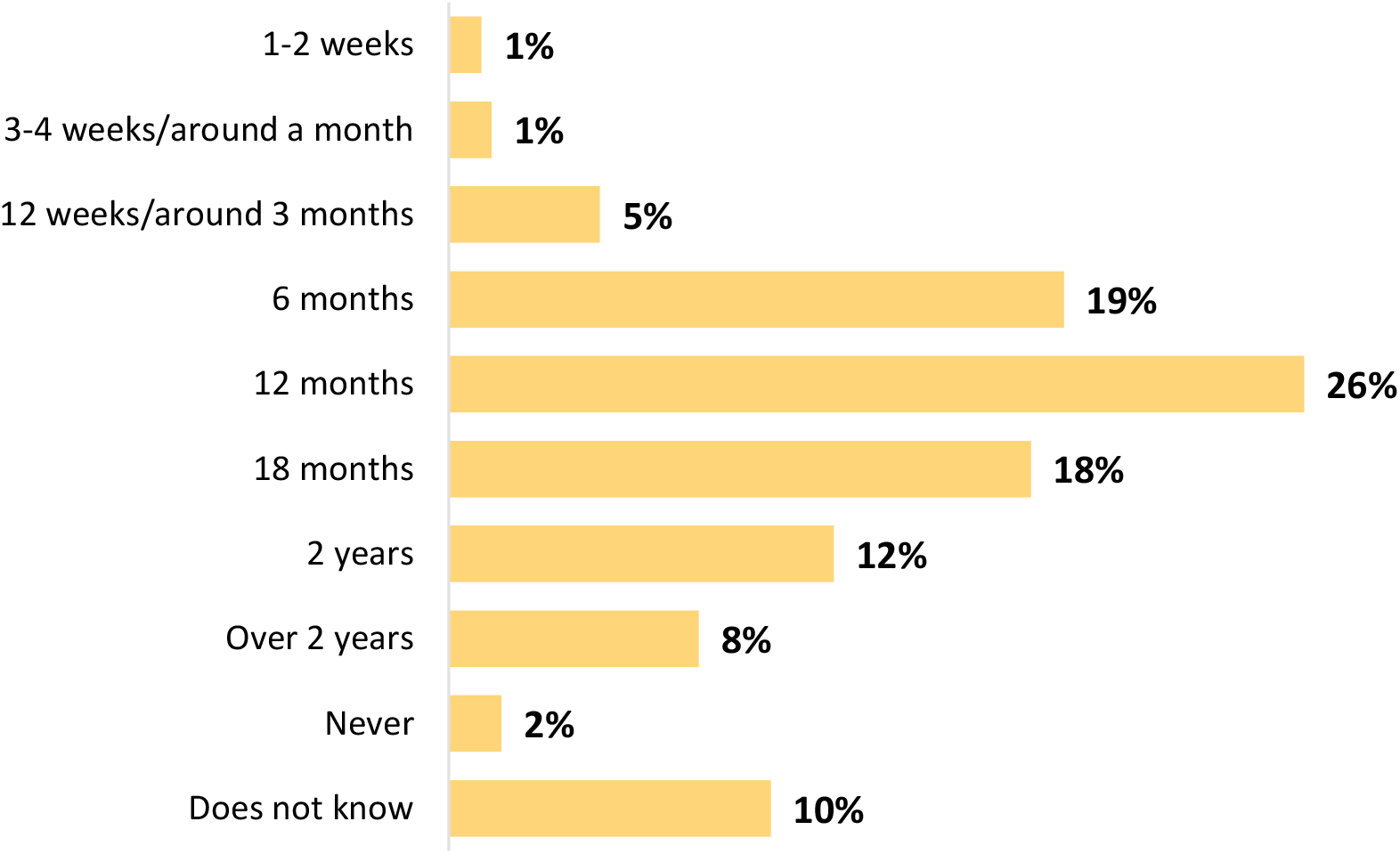
People’s Beliefs about COVID-19 Vaccination, Colombia 2020.

**Figure 21.**
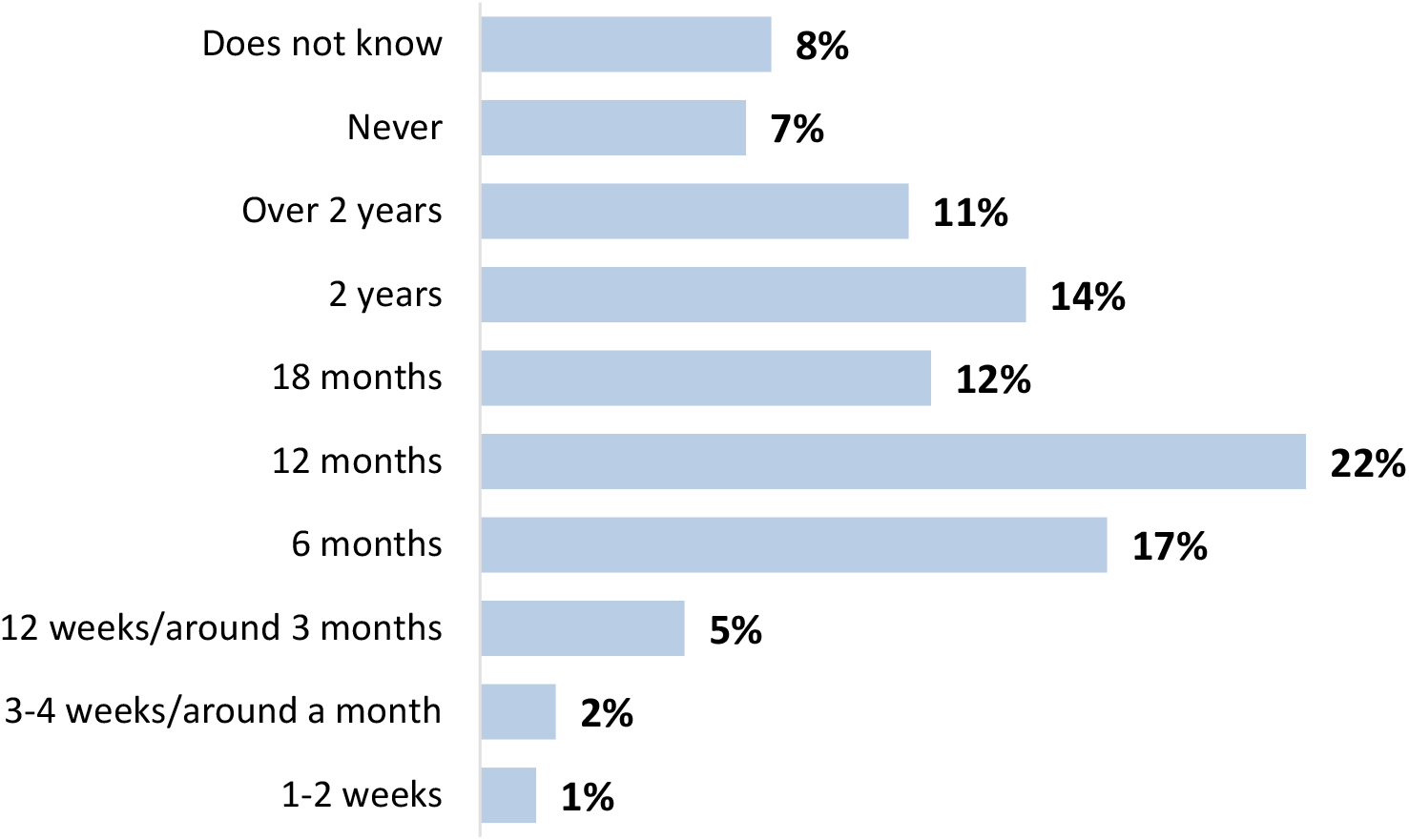
Life Will Return to “Normal” (Living Like They Used To), Colombia 2020.

### 11) Community Engagement

43% of respondents mention that measures have been taken or campaigns conducted in their neighborhood, district or municipality to prevent the spread of COVID-19.

Among the measures taken in neighborhoods, districts, or municipalities to prevent the spread of COVID-19 are:

- 91% restricted entrance to supermarkets and shopping centers by the ID restriction measure (pico y cédula)
- 70% use of protective gear by guards and general services staff
- 60% key messages on safe behavior in public spaces in neighborhoods
- 54% access restriction for delivery staff in apartment and home complexes.
- 53% cleaning and disinfection of streets, avenues and inside apartment and home complexes
- 50% safety and biosecurity campaigns disseminated through social media
- 43% delivery services within the neighborhood or community
- 31% closure of access roads to municipalities that draw tourism
- 29% identification which population are at higher risk
- 24% community mapping to identify critical areas
- 19% arrangement of public spaces for delivery of cleaning and hygiene products

Reported ways in which the community engaged:

- 31% helping to decide what information is shared with the public and how it is communicated
- 27% reviewing ideas and contributing to decision making
- 25% supporting local communities in response to the outbreak
- 7% doing live events online
- 6% sharing personal experiences with the virus
- 38% were not interested in participating

42% of young people (18-34 years of age) are interested in participating in community activities, but the interest in participating in these types of activities decreases with age.

### 12) Changes in Immediate Expectations

Changes that people would accept in the case that no vaccine or treatment becomes available in the short or medium term:

#### Children’s Education

82% of respondents agree with most children remaining home schooled, 85% agree that parents should have the ability to choose whether or not to send their children to school, and 17% agree that parents must send their children to school when the government instructs it.

- Among younger people (18-24 years) we find the lowest percentages of agreement (76%) for children continuing to be educated at home and for parents to choose whether to send their children to school.
- Older people (50-54 years) show the highest percentage of agreement (32%) with parents having to send their children to school when the government instructs it.
- Women agree at higher percentages with children continuing to be educated at home (83%).
- 22% of men accept that parents must send their children to school when the government instructs it, while 15% of women accept the same notion.
- 83% of those who adhered mandatory isolation agreed with children continuing to be educated from home.

#### Work from home

86% agree with employees being able to choose whether to work at their office or from home, and 27% accept that employees would have to go to their place of work when the government instructs it.

- No differences were found among the opinions of respondents by age and gender for either situation.
- 91% of the people who are working from home agree that employees should have the ability to choose where they work.
- 80% of the women who are heads of households, and 88% of the people who adopted mandatory isolation agree that employees should be able to choose whether to work from home or at the office.

#### Daily life

82% agree with the notion that no major sporting or cultural events take place in front of a live audience.

- Among younger people (18 to 24 years old) we found the lowest levels of acceptance; the level of acceptance increased with age.
- No differences in opinion by gender were found.
- 84% of people who adopted mandatory isolation agree with not allowing sporting or cultural events to take place.

56% agree with people deciding by their own accord whether to attend live sporting or cultural events.

- Among older people (50 years of age and over) we found the lowest levels of acceptance, while in younger people the level of acceptance increased.
- No differences by gender were evident.
- 52% of people who adopted mandatory isolation agree with people deciding by on their own accord whether to attend sports or cultural events.

95% agree with people being forced to wear face masks when they leave their homes.

- There are no differences by age and gender and acceptance levels were above 90%.
- 96% of people who adopted mandatory isolation agree with people being forced to wear face masks.

86% do not agree with people deciding whether or not to use face masks when they leave home.

- The highest levels of disagreement (94%) were found among people aged 50-54 years.
- There were no differences in opinion by gender.
- 88% of people who adopted mandatory isolation do not agree with people deciding whether or not to use face masks when they leave their homes.

37% of people agree with young people having fewer restrictions in their activities than older people, as they face a lower risk of contracting COVID-19.

- The highest levels of approval (45%) were found amongst younger people (18 to 24 years old).
- People aged 60 and older reported the lowest levels of approval (24%) to the notion of young people having fewer restrictions.
- No differences were found across genders.
- 35% of the people who adopted mandatory isolation agree with young people having fewer restrictions than older people.

90% agree with having tighter restrictions in neighborhoods, towns, or municipalities that experience outbreaks than national restrictions, and 85% agree with their neighborhood, district, or municipality having tighter restrictions.

- The lowest levels of approval are found among young people both for tight restrictions in neighborhoods, districts, or municipalities in general as well as having higher restrictions in their neighborhood, district, or municipality.
- People aged 45-49 present the highest levels of acceptance to strict restrictions both for neighborhoods, districts, or municipalities in general as well as for their own neighborhoods, districts, or communities.
- No differences were found by gender.

#### Financial and Work Expectations

27% believe they are likely to lose their jobs as a result of COVID-19, 15% have already lost their jobs and 9% are already experiencing financial difficulties.

- Younger people (18-29 years of age) have the highest rates of job loss and financial hardship.
- 37% of people aged 50-54 and 34% of people aged 40-44 believe they are likely to lose their jobs.
- 35% of men believe they are likely to lose their job and 57% believe they are likely to have financial difficulties.
- 10% of women have already experienced financial difficulties.
- 29% of people who are going to work and 25% of those working from home believe they are likely to lose their jobs.
- 54% of people who are going to work and 43% of those working from home believe they are likely to experience financial difficulties.
- 33% of female heads of household believe they are likely to lose their job and more than half (55%) believe they will experience financial difficulties.

#### Expectations Regarding Vaccines

2% of respondents think they will never be able to get a vaccine against the virus, 19% believe they will get it in two years or more, 44% within 12 or 18 months, and 26% within six months or less.

- 32% of younger people (18-24 years of age) think they will be able to be vaccinated within six months or less and 53% of people aged 45-49 believe they will be able to get vaccinated in 12-18 months.
- 47% of men and 43% of women think they can be vaccinated within 12 to 18 months.
- 45% of people who have adopted mandatory isolation think they can get vaccinated within 12 to 18 months.

#### Return to Normalcy

54% of people think that all children will return to elementary school in six months, 32% believe this could happen within 12 to 18 months, and 5% within two years or more. 58% think that all children will return to high-school in six months, 28% in 12 to 18 months, and 4% in two years or more.

7% of people think that life will never return to “normal” in a context in which people live like they used to, 26% believe that this will happen in two years or more, 34% within 12 or 18 months, and 6% within six months or less

### Key Messages

1. Protective measures such as wearing face masks and avoiding going out to social events increased. People who had to go out during quarantine did so mainly to: go to work, look for work, or due issues with anxiety or depression.
2. Changes in people’s sexual lives were reported, and it was possible to evidence a drop in basic access to health services such as services related to access to contraceptives and specialized medical visits such as gynecology and endocrinology. These explain certain unmet needs in sexual and reproductive health as reported, which, in turn, are explained by the increase in barriers to access these specific services. Among the most frequently reported barriers, we found: delays in authorization for health care services (by health care providers), lack of financial resources, lack of information such as people not knowing that the service was available, and peopleś fear of going out.
3. Mental health issues (feeling fatigued and hopeless) and worries (the pandemic not being controlled and the vaccine not reaching the country soon) increased during quarantine. Most people have experienced mental health problems, especially young female heads of household (39 years of age). Living in quarantine increased low self-esteem, fatigue, feelings of fear or hopelessness, as well as anxiety and stress compared to the survey performed in April.
4. Regarding expectations, some people say they would accept significant changes in the long term if a vaccine or treatment does not become readily available: they agree that children should continue being educated from home; that employees should be able to choose whether to work from home or at the office; that the use of face masks should be made mandatory and that targeted quarantines should be implemented in areas with a high number of cases. More than half of the people think they will be able to get the COVID-19 vaccine within six months or more, and they also think that life will return to normal in a year or more.

## Data Availability

This database is freely accessible and for research purposes. If you use it, please acknowledge the Profamilia Association for the credit and authorship of the collection, storage and debugging of your work and material produced.
http://profamilia.org.co/wp-content/uploads/2020/11/BD-DEFINITIVA-SOLIDARIDAD-II.sav
https://profamilia.org.co/wp-content/uploads/2020/11/Diccionario-de-datos-Solidaridad-II.pdf

## Acknowledgments

This study was funded by Profamilia Association.

